# Consumption of legumes and risk of hepatobiliary diseases among humans aged 2+ years: a scoping review

**DOI:** 10.1101/2024.11.15.24317376

**Authors:** Fie Langmann, Christopher Fisker Jensen, Vibeke Lie Mortensen, Line Rosendal, Niels Bock, Christina C. Dahm

## Abstract

**Objectives:** To identify and map the literature regarding human consumption of legumes and potential relationships with hepatobiliary diseases.

**Background:** Consuming legumes might reduce lipid accumulation in the liver while potentially impeding the functionality of the bile duct and gallbladder. As dietary trends shift toward promoting legume consumption, exploring the positive and negative impacts on hepatobiliary health becomes crucial.

**Methods:** This scoping review explored the intake of dietary pulses and legumes (excluding broader dietary patterns) and their relation to hepatobiliary diseases like non-alcoholic fatty liver disease, gallstones, and gallbladder removal among individuals above the age of two years. The review included textbooks, expert opinions, and searches in four databases (PubMed, CINAHL Complete, Embase, and Web of Science). Two reviewers extracted data independently from each article. The synthesis of results was presented narratively by hepatobiliary outcomes. Unpublished studies and grey literature sources were sought out.

**Results:** From 19,881 records retrieved, 29 were included - 17 on non-alcoholic fatty liver disease and 12 on gallbladder diseases. Most studies were observational, but a few were narrative reviews. Some studies found a protective association between legume consumption and non-alcoholic liver disease, while others found no association. Overall, there was no clear association between legume consumption and gallbladder diseases. The studies varied in methodological quality, and confidence in the findings was low.

**Conclusion:** The association between legume consumption and non-alcoholic fatty liver disease was inverse or non-significant. The association between legume consumption and gallbladder disease was inconclusive. Further research is needed to draw firmer conclusions. **Keywords**: Dietary pulses; Gallstone; Hepatobiliary health; Legumes; Non-alcoholic fatty liver disease

## Introduction

Non-alcoholic fatty liver disease (NAFLD) is a chronic liver disease, anticipated to affect around 25% of adults worldwide by 2026. ^1^ It arises from excessive fat accumulation in the liver, unrelated to notable alcohol consumption, parenteral nutrition, or medication use. ^2^ The disease encompasses non-alcoholic fatty liver (NAFL) characterized by ≥5% hepatic fat without apparent damage, and non-alcoholic steatohepatitis (NASH) characterized by ≥5% hepatic fat with inflammation, with or without fibrosis. ^2^ Diagnosis with NAFLD is strongly associated with metabolic syndrome (MetS) and elevated risk of developing liver cancer compared to those without the condition. ^2,3^ No approved drug therapy exists for NAFLD, and lifestyle modifications, notably dietary changes, form primary clinical recommendations. Diets emphasizing plant-based protein sources and high dietary fiber contents are recommended. ^2^

Compared to meats, legumes offer rich protein content, low saturated fat, energy density, and abundant dietary fiber, all crucial components for a healthy and balanced diet. ^4–6^ Diets enriched with legumes have further been linked to overall better diet quality and health outcomes. ^7^ Pulses (or grain legumes) are a subtype of legumes referring to crops harvested for dry grain such as lentils, chickpeas, and beans, but not fresh legumes like green beans. The term *legumes* is often used as an umbrella term for both pulses and fresh legumes. ^4^ Legumes constitute fundamental elements of various plant-based diets, providing health advantages and reducing the risk of lifestyle-related diseases compared to Western diets high in red meats, fats, and sugars. ^8–10^

Prior research has shown an advantageous impact on NAFLD of dietary patterns integrating legumes, such as the Mediterranean and vegetarian diets. These benefits are often associated with the abundant presence of bioactive compounds like polyphenols and antioxidants, and the high fiber content of legumes. ^11,12^ Although legumes boast substantial amounts of bioactive components and fibers, ^13^ these bioactive components are also present in various other food groups. There is a gap in understanding the comprehensive impact of whole grain legume consumption or processed legumes used in food products like plant-based meat alternatives.

Elevated legume intake has been linked to inhibited digestion of fats and carbohydrates, ^14,15^ which might mitigate excessive fat accumulation in hepatocytes. ^16,17^ Consequently, a lower risk of NAFLD could ensue with increased legume consumption. While human and animal studies have demonstrated the hepatobiliary benefits of plant-based diets, ^12,18^ research on legumes in isolation is scarce, rendering the evidence indirect and restricted. The pathways of NAFLD involve the bile duct and gallbladder function, where excess lipid fractions are excreted through the gallbladder. ^19^ Polyphenols, peptides, and other leguminous compounds have been associated with increased cholesterol synthesis and biliary cholesterol secretion. ^14,15,20^ This may result in cholesterol saturation within the bile, potentially raising the likelihood of gallstone development, ^15,20,21^ and complications like gallbladder inflammation. ^22,23^ Following symptom onset, the annual risk of gallbladder removal is 1-2%.^24^ Greater legume consumption might therefore correlate with a higher risk of gallstones, through the pathways that concurrently minimize the risk of NAFLD. ^19,20,25^

With greater emphasis on reducing the environmental impact of Western diets through consumption of legumes, ^5^ a comprehensive investigation of potential adverse and beneficial effects is important. This scoping review therefore aimed to examine the breadth of literature about the consumption of legumes and dietary pulses, whether as whole grains or components within foods such as plant-based meat alternatives, in relation to NAFLD, gallstone, and gallstone-related diseases. This review focused on legume crops usually consumed as protein sources instead of meats, such as pulses, while excluding peanuts and legume crops used for sowing forage, such as clovers. ^4^ It targets individuals aged 2+ years, omitting young children recommended to breastfeed for at least 2 years, ^26^ without prior hepatobiliary conditions or limitations affecting nutrient digestion or absorption, across diverse geographical regions. While previous research has delved into legume components, such as polyphenols or antioxidants, this scoping review explores the implications of consuming legumes beyond clinical contexts.

A preliminary search of PROSPERO, the Cochrane Database of Systematic Reviews, and *JBI Evidence Synthesis* was conducted. No current or in-progress scoping reviews or systematic reviews on the topic were identified.

## Review questions

What research has been reported on whole grain legumes or products based on whole grain legumes and hepatobiliary disease in humans without previous hepatobiliary disease?

What evidence of the association between legume consumption and hepatobiliary diseases has been reported for children, the general adult population, adults with cancers, and adults with cardiovascular diseases?

What hepatobiliary mechanisms is the consumption of whole grain legumes considered to affect?

## Inclusion criteria

### Participants

This review encompassed studies involving individuals aged 2 years or older with no prior diagnosis of NAFLD, NASH, metabolic dysfunction-associated fatty liver disease (MAFLD), metabolic dysfunction-associated steatotic liver disease (MASLD), metabolic dysfunction-associated steatohepatitis (MASH), gallstone (cholelithiasis), or gallbladder inflammation (cholecystitis or biliary cholangitis). The review protocol outlined that literature involving individuals who had their gallbladder removed (cholecystectomy) were not eligible for inclusion. ^27^ Cholecystectomy is, however, a common treatment and proxy for gallstones, ^28,29^ and studies investigating this outcome were therefore included in the literature search. Literature involving individuals who underwent partial or full surgical removal of the liver or gallbladder before entry to the study was not considered. Additionally, studies involving individuals with gastrointestinal conditions impeding digestion or nutrient absorption, such as ileostomy, colostomy, Crohn’s Disease, etc., were excluded. The review aimed to comprehensively map and describe the literature irrespective of sex. Articles focusing on animal studies were not included.

### Concept

This review focused on studies examining the consumption of whole-grain legumes or products derived from whole-grain legumes, such as meat-substitute products, and their association with non-cancerous hepatobiliary diseases.

Legumes were defined culinarily, encompassing peas, lentils, and beans (excluding coffee and cacao beans). Fatty legumes like peanuts were not included. Studies of specific legume components (e.g., specific antioxidants, fibers, or peptides) were not within the scope of this review. Additionally, literature about *Leguminosae* or *Fabaceae* not consumed as whole grains (e.g., stems, leaves, or flowers) was not included. Materials exploring legumes within enteral or parenteral nutrition contexts were beyond the scope of this review. The outcomes of the included literature were NAFLD, NASH, MAFLD, MASLD, MASH, gallstones, gallbladder inflammation, bile duct obstruction, and gallbladder removal. In instances where the hepatobiliary condition was present at the study baseline, only investigations of NAFLD progression to NASH or case-control studies retrospectively assessing legume consumption as an exposure were eligible. In 2023, the European, American, and Latin American Associations for the Study of the Liver, announced the change of NAFLD to MASLD and NASH to MASH. ^30^ The change was made to avoid stigmatization of affected individuals and to classify the disease on the presence of specific characteristics rather than the non-presence of alcohol. ^30,31^ This change in nomenclature was accepted after the review process started and thus incorporated into the literature search of this review.

### Context

Legumes and dietary pulses are consumed across various populations, cultures, and settings, serving as a dietary staple in many developing countries. As such, the review context encompassed this diverse array of settings, also including trial settings, where an intervention was evaluated. To ensure a comprehensive mapping of available evidence, restrictions based on country or date were not imposed. Articles published in Danish, Swedish, Norwegian, and English were considered for inclusion. Of the four languages, the literature search exclusively identified articles composed in English. The literature search included all literature up until June 30^th^, 2023.

### Types of sources

This scoping review considered all types of quantitative and qualitative study designs and reviews for inclusion. Quantitative studies, whether randomized or non-randomized controlled trials or observational, were included. Eligible qualitative studies included phenomenology, grounded theory, ethnography, and thematic analysis. Literature without evidence level that met the inclusion criteria, along with text and opinion papers, was also considered for the scoping review. Any peer-reviewed or expert publications of consensus guidelines were eligible for inclusion.

## Methods

This scoping review was conducted in line with the Preferred Reporting Items for Systematic Reviews and Meta-Analyses extension for Scoping Reviews (PRISMA-ScR) ^32^ following an *a priori* protocol. ^27^

### Search strategy

A 3-step search strategy was performed to locate both published and unpublished literature. In step 1, an initial limited search of PubMed (National Center for Biotechnology Information, MD USA) was undertaken on February 17^th^, 2023. The text words contained in the titles and abstracts of relevant articles, and the index terms used to describe the articles were used to develop a full search strategy. In step 2, the full search strategy, including all identified keywords and index terms was adapted for each information source and undertaken on July 24^th^, 2023. Fatty liver diseases have several different clinical and histological characteristics and varying nomenclature including NAFLD, MAFLD, and MASLD, which has changed over time. ^30,31^ Being observant of the recent change in nomenclature for NAFLD, ^33,34^ the literature search was also conducted for MASLD, MAFLD, and MASH until June 30^th^, 2023. The full search strategies are provided in Appendix I and II. In step 3, the reference lists of articles selected for full-text review were screened for additional papers. No relevant literature on MAFLD, MASLD, or MASH was found, and this review will use the terms NAFLD and NASH throughout the text. All databases were searched from the date of inception until June 30^th^, 2023, apart from sources of unpublished and grey literature, which were narrowed to only include literature published in the year preceding June 30^th^, 2023. This decision was made to prevent excessive screening time.

The databases searched included PubMed (National Center for Biotechnology Information, MD USA), Embase (Elsevier B.V., Amsterdam, Netherlands), CINAHL Complete (EBSCO Information Services, MA, USA), and Web of Science (Clarivate Analytics, PA, USA). Sources of unpublished studies and gray literature searched included Google Scholar (Google Inc., CA, USA) and ProQuest (ProQuest LLC, MI, USA). OpenGrey was discontinued in 2020 and therefore not included as a literature source. ^35^

### Study selection

All identified records were collated in Covidence (Veritas Health Innovation Ltd, Melbourne, Australia), and duplicates were removed. Following a pilot test, titles and abstracts were screened by two independent reviewers for assessment against the inclusion and exclusion criteria for the review. Articles not meeting the criteria for inclusion were excluded, and potentially relevant papers were retrieved in full and assessed in detail against the inclusion criteria by two independent reviewers (FL and VLM, LR, CFJ, or NB). Full-text studies that did not meet the inclusion criteria were excluded, and reasons for their exclusion are provided in Appendix III. Disagreements between reviewers at any stage of the selection process were resolved through discussion or with a third reviewer (CCD).

### Data extraction

Data were extracted into a data extraction table based on the draft form presented in the protocol. ^27^ Data extraction from the included literature was carried out by one author (FL) and reviewed by a second author (VLM, CFJ, LR, NB, or CCD). The data extraction table was piloted by two reviewers (FL and CCD) and minimally modified to include information on the author, aim, and study type in the same column. Extracted data included specific details about the population, concept, context, statistical methods, confounding factors, and key findings. An outline of the final data extraction table used for data extraction can be found in Appendix IV. Any disagreements between reviewers were resolved through discussion or with a third reviewer.

### Data analysis and presentation

Summaries of all included studies are presented in tables according to hepatobiliary disease type (NAFLD or gallbladder diseases [GBD]) and further presented by i) study design for evidence-level articles, or ii) article type for non-evidence articles. Results from evidence-level articles are presented in tables developed from the draft results table presented in the protocol for this review^27^ and summarized in narrative reviews together with data from non-evidence articles to answer the review questions. All evidence-level articles were subject to a quality assessment to ensure that any conclusions, and implications for research or practice, considered not only results but also the methodological soundness of the included studies. Two independent reviewers (FL and VLM, CFJ, LR, or NB) assessed methodological quality following assessment templates (Appendix V) for the specific study design.

Cohort and case-control studies were assessed with the Scottish Intercollegiate Guidelines Network (SIGN) checklists for observational studies. ^36^ SIGN includes 14 statements for cohort studies evaluating the internal validity followed by an overall evaluation with three statements and a note section. Case-control studies were assessed based on 11 statements evaluating the internal validity followed by an overall evaluation with three statements and a note section. For both cohort and case-control studies, each statement was answered yes, no, or unknown and the overall evaluation of minimized bias was rated as high, acceptable, or unacceptable based on the internal validity and consensus reached between the assessors. The SIGN checklist for case-control studies included an unclear description for assessing statement, 1.9: “Exposure status is measured in a standard, valid, and reliable way.” The description focused solely on the measurement of outcomes rather than exposure status. ^36^ SIGN was contacted to clarify the statement and sent a response to the authors on March 1^st^, 2024. Due to the late response, the authors reached a consensus in November 2023 that the statement should include exposure and outcome measurement assessments, as both are essential for the methodological quality of case-control studies. Statement 1.9 was thus amended to: “Exposure and outcome status are measured in standard, valid, and reliable ways”. With the assessment description: “The study needs to outline the exposure and outcome measures employed, explicitly. It should detail how exposure levels were determined, ensuring a clear understanding of whether participants were exposed to the investigated factor and to what extent. Additionally, if the outcomes involve any subjective elements, the study should offer evidence validating the reliability of these measures before their application. Clear, comprehensive descriptions of reliable measures enhance the study’s credibility. Studies omitting outcome measures or heavily rely on secondary outcomes may warrant rejection”.

Cross-sectional studies were assessed using the Appraisal of Cross-sectional Studies (AXIS) tool comprising 18 statements evaluating internal validity. Two statements were used to evaluate ethics and conflicts of interest. ^37^ The reviewers added a note section to the assessment tool. Each statement was answered yes, no, or unknown and the overall methodological quality was evaluated as high, moderate, low, or very low based on consensus reached between reviewers.

Systematic reviews and meta-analyses were assessed with A MeaSurement Tool to Assess systematic Reviews (AMSTAR-2) based on 16 statements that answered yes, no, or partly yes. ^38^ The overall confidence in results was evaluated as high, moderate, low, or critically low based on consensus reached between reviewers.

## Results

### Article inclusion

The literature search of published literature yielded 19,881 records. After the removal of duplicates, 16,498 titles and abstracts were screened, and 57 full-text records were assessed for eligibility with 33 not meeting the inclusion criteria (Appendix III). Five records were included through other methods including hand searches of reference lists of articles assessed for full-text eligibility, and from the search of unpublished and grey literature. Twentynine records were included in the review (Figure 1).

**Figure 1:**
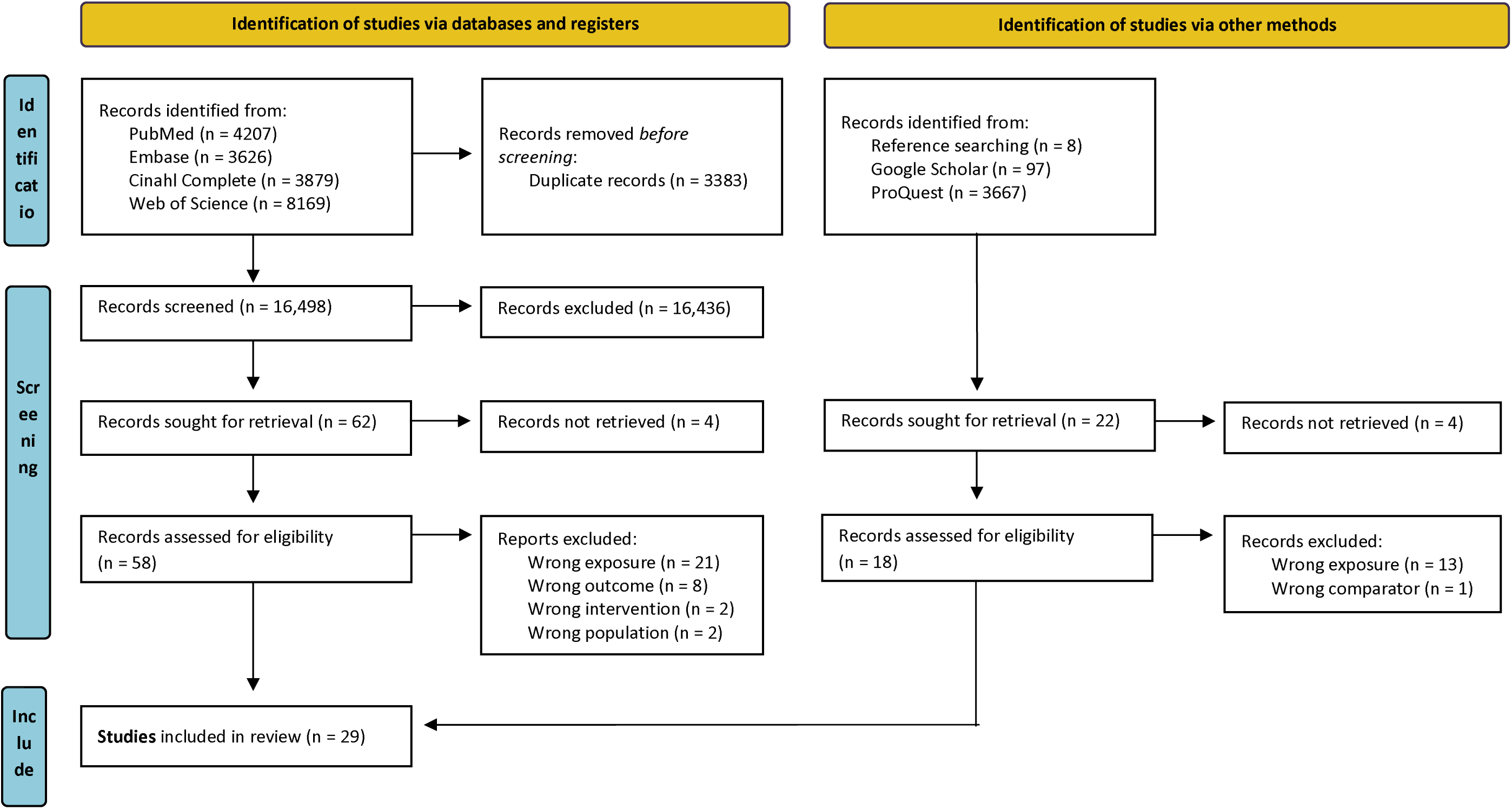
Search results, study selection and inclusion process following the PRISMA Extension for Scoping Reviews. ^32^

### Characteristics of included studies

Of the 29 articles included, two presented data from the same study, ^39,40^ the narrative reviews presented data from four primary studies^25,41–43^ and one systematic review, ^3^ which was also included in the scoping review. The systematic review presented data from four primary studies. ^42–45^

The characteristics of the included studies are summarized in Table 1 and elaborated in Appendix VI. There were 17 articles focusing on NAFLD (1 cohort study, ^40^ 5 case-control studies, ^43,46–49^ 9 cross-sectional studies, ^39,42,44,45,50–54^ 1 systematic review, ^3^ 1 narrative review^55^). Most were from Asian countries (5 China, 1 Korea, 1 Taiwan 1 Japan) or South European countries (4 Italy, 1 Spain) with the remaining from Western Europe (1 Germany, 1 UK) and the Middle East (2 Iran). Twelve articles focused on GBD (2 cohort studies, ^41,56^ 5 case-control studies, ^25,57–60^ 2 cross-sectional studies, ^61,62^ 3 narrative reviews^21,63,64^). The studies were geographically spread across Asia (2 India, 1 Pakistan), South Europe (1 France, 1 Greece, 1 Italy), North America (3 USA), Western Europe (1 The Netherlands), Middle East (1 Saudi Arabia) and one of unknown origin. ^64^

**Table 1.** Overview of eligible research study characteristics.

| Study design | Number and characteristics | Populations included | Exposure | Outcomes |
| --- | --- | --- | --- | --- |
| Articles focusing on NAFLD<sup>a</sup> |  |  |  |  |
| Cohort studies<sup>40</sup> | 1 study published in 2023. Number of participants: 15,838. China. | Healthy general population without CVD<sup>b</sup> or cancer. | Replacing 25g of other foods with legumes. | NAFLD |
| Case-control studies<sup>43,46-49</sup> | 5 studies published in 2014-2021. Number of participants: 210-4091. 2 Iran, 1 Italy, 1 Korea, 1 China. | Healthy general population with low consumption of alcohol and without hepatitis B or C, medically induced fatty liver, liver dysfunction, CVD, endocrine disorders, or cancers. | Legume intake in frequencies, tertiles, g/day, or servings/week. | NAFLD |
| Cross-sectional studies<sup>39,42,44,45,50-54</sup> | 9 studies published in 2015-2022. Number of participants: 136-24,622. 2 Italy, 2 China, 1 Spain, 1 Taiwan, 1 Germany, 1 Japan, 1 UK. | High CVD-risk individuals and healthy general population with low consumption of alcohol and tobacco, and without hepatitis, medically induced fatty liver, liver dysfunction, CVD, cancer, or endocrine or GI<sup>c</sup> disorders. | Weekly servings of legumes, legume intake (tertiles or g/day), or replacing 7g meat protein with soy. | NAFLD, NAFLD severity, liver fibrosis, or liver stiffness. |
| Systematic | 1 review published in 2020. Included | Adult non-pregnant participants with low | Legume intake. | NAFLD |
| reviews<sup>3</sup> | 4 articles within the scope of this review (1 case-control, 3 cross-sectional). 1 China. | alcohol consumption and without hepatitis B or C, HIV<sup>d</sup>, IBD<sup>e</sup>, coeliac disease, or autoimmune hepatitis. |  |  |
| Narrative reviews<sup>55</sup> | 1 article published in 2021. 1 Italy. | Mixed populations, healthy adults, and NAFLD patients. | Legume intake. | NAFLD |
| Articles focusing on gallbladder diseases |  |  |  |  |
| Cohort studies<sup>41,56</sup> | 2 studies published in 2006-2017.<br>Number of participants: 64,052-77,090. 1 France, 1 USA. | Women from E3N<sup>f</sup> or Nurses' Health Study without cancer, gallstone, or cholecystectomy. | Legume intake g/day or servings/week. | Cholecystectomy |
| Case-control studies<sup>25,57-60</sup> | 5 studies published in 1990-2020.<br>Number of participants: 200-819. 1 India, 1 Pakistan, 1 Greece, 1 Saudi Arabia, 1 The Netherlands. | Cholecystectomy and gallstone cases matched with healthy controls. Included individuals not pregnant and without cancer, hepatitis, or AIDS<sup>g</sup>. | Daily, weekly, and monthly legume intake. | Gallstone |
| Cross-sectional studies<sup>61,62</sup> | 2 studies published in 2000-2010.<br>Number of participants: 4641-11,392. 1 USA, 1 India. | Mexican Americans and Indians with gallstone matched with healthy adults | Intake of chickpeas (yes, no) or beans (frequency). | GBD<sup>h</sup> |
| Narrative reviews<sup>21,63,64</sup> | 3 articles published in 1989-2019. 1 Italy, 1 USA, 1 unknown. | Populations consuming large amounts of legumes | Legume intake. | Gallstone, biliary cholesterol, GBD |

<sup>a</sup>NAFLD, non-alcoholic fatty liver disease. <sup>b</sup>CVD, cardiovascular disease. <sup>c</sup>GI, gastrointestinal. <sup>d</sup>HIV, human immunodeficiency virus. <sup>e</sup>IBD, inflammatory bowel disease. <sup>f</sup>E3N, Etude Epidémiologique auprès de Femmes de la Mutuelle Générale de l'Education Nationale. <sup>g</sup>AIDS, acquired immunodeficiency syndrome. <sup>h</sup>GBD, gallbladder disease.

### Review findings

The literature search did not return any studies on children or individuals with cancers. As only two studies investigated NAFLD among individuals with CVD, the results are presented together with the studies investigating NAFLD among healthy individuals. There were no GBD studies on individuals with CVD. Table 2 and 3 presents results and quality assessment ratings for studies focusing on NAFLD and GBD, respectively.

**Table 2.** Results table for studies investigating legume consumption and NAFLD^a^, including quality assessment rating.

| Author | Population | Exposure | Outcome | Result<sup>b</sup> | Quality<sup>c</sup> |
| --- | --- | --- | --- | --- | --- |
| Cohort studies |  |  |  |  |  |
| Zhang et al.<br>(2023)<sup>40</sup> | TCLSIH<sup>d</sup>, n=14,541, 59.9% females, median age 35.9 years.<br>GNHS<sup>e</sup>, n=1297, 69.5% females, median age 59.7 years. China. | Replacing 25 g legumes with<br>equal mass of other foods. | NAFLD | ≤ | High |
| Case-control studies |  |  |  |  |  |
| Bahrami et al.<br>(2019)<sup>43</sup> | 196 NAFLD cases, 803 age and sex-matched controls, 51%<br>females, mean age 41.9 years. Iran. | Weekly servings of legumes. | NAFLD | < | High |
| Giraldi et al.<br>(2020)<sup>46</sup> | 371 NAFLD cases, 32.8% female, mean age 59 years. 444 controls,<br>41.2% female, mean age 45 years. Italy. | Low or high legume intake. | NAFLD | < | Acceptable |
| Han et al.<br>(2014)<sup>47</sup> | 169 NAFLD cases, 179 controls, 52.3% females, age 20-69 years.<br>Korea | Tertiles of bean intake<br>(g/day). | NAFLD | = | Acceptable |
| Hao et al.<br>(2021)<sup>48</sup> | 2602 NAFLD cases, 1447 genotype-matched controls, 69-71%<br>females, mean age for cases: 53 years, controls: 53.1 years. China. | Weekly consumption<br>frequency of beans. | NAFLD | < | Unacceptable |
| Tutunchi et al.<br>(2021)<sup>49</sup> | 105 NAFLD cases, 105 age and sex-matched controls, 57.2%<br>females, mean for age cases: 45.6 years, controls: 45.4 years. Iran. | Tertiles of legume intake<br>(g/day). | NAFLD | = | Unacceptable |
| Cross-sectional studies |  |  |  |  |  |
| Baratta et al. (2017) <sup>44</sup> | 584 patients with cardiovascular risk factors, 38.2% females, mean age 56.2 years. Italy. | Weekly servings of legumes <2 or ≥ 2. | NAFLD | = | Low |
| Bullón-Vela et al. (2019) <sup>50</sup> | 328 individuals with MetS<sup>f</sup>, 45.1% females, mean age 65.8 years. Spain. | Tertiles of legume intake (g/day). | NAFLD | < | Moderate |
| Chan et al. (2015) <sup>45</sup> | 797 adults, 58.3% females, mean age 48.1 years. China. | Tertiles of soy food intake (g/day). | NAFLD | = | Very low |
| Chiu et al. (2018) <sup>42</sup> | 4625 participants from the Tzu Chi Health Study. Vegetarian: mean age 54 years, 59% females. Non-vegetarian: mean age 55 years, 78% females. Taiwan. | Replacing 7g legume protein with equal mass protein from other foods. | NAFLD | = | Low |
| Mirizzi et al. (2019) <sup>51</sup> | 136 individuals from NUTRIATT-RCT<sup>g</sup>, 42% females, mean age 49.6 years. Italy. | Legume intake (g/day). | NAFLD | ≤ | Moderate |
| Vijay et al. (2022) <sup>52</sup> | 993 NAFLD cases and 973 controls, from the Trivandrum NAFLD cohort, cases: 54.4%, female, mean age 48.16 years, controls: 67.5% females, mean age 45.90 years. India. | Legume intake (g/kg/day). | Degree of liver fibrosis | < | Moderate |
| Watzinger et | 136 overweight individuals from the HELENA trial, without | Legume intake (g/day). | NAFLD | = | Moderate |
| al. (2020)<sup>53</sup> | NAFLD: 57.8 %, females, mean age 49.8 years, with NAFLD: 44.4% females, mean age 50.1 years. Germany. |  |  |  |  |
| Yabe et al. (2021)<sup>54</sup> | 248 with NAFLD, 52.4% females, mean age 55.8 years. 101 without NAFLD, 71.3% females, mean age 58.4 years. Japan. | Weekly frequency of soy and soybean intake. | Liver fibrosis | < | Low |
| Zhang et al. (2020)<sup>39</sup> | 24,622 participants from TCLSIH. Without NAFLD: 58.8% females, mean age: 38.4 years. With NAFLD: 27.7% females, mean age: 42.4 years. China. | Weekly frequency of soy and soybean intake. | NAFLD | ≤ | High |
| Systematic review |  |  |  |  |  |
| He et al. (2020)<sup>3</sup> | Of 24 included studies, four investigated legume intake and NAFLD. Inclusion: Adults, observational studies investigating food groups and validly assessed NAFLD. | Food groups - legumes composed one food group. | NAFLD | ≤ | Moderate |

<sup>a</sup>NAFLD, non-alcoholic fatty liver disease. <sup>b</sup>Results are presented as “<” corresponding to an inverse association, “≤” corresponding to an inverse or no association, and “=” corresponding to no association. <sup>c</sup>Quality refers to the methodological quality assessment of the study. <sup>d</sup>TCLSIH, Tianjin Chronic Low-grade Systemic Inflammation and Health Cohort. <sup>e</sup>GNHS, Guangzhou Nutrition and Health Study. <sup>f</sup>MetS, metabolic syndrome. <sup>g</sup>NUTRIATT-RCT, Nutrition and Activity Randomized Controlled Trial.

**Table 3.** Results table for studies investigating legume consumption and gallbladder disease, including quality assessment rating.

| Author | Population | Exposure | Outcome | Result<sup>a</sup> | Quality<sup>b</sup> |
| --- | --- | --- | --- | --- | --- |
| Cohort studies |  |  |  |  |  |
| Barré et al.<br>(2017)<sup>56</sup> | 64,052 women from the E3N<sup>c</sup> cohort, mean age 53.7 years for those reporting the outcome, 52.6 years for those not reporting the outcome. France. | Legume intake (g/day) | Self-reported cholecystectomy | < | High |
| Tsai et al.<br>(2006)<sup>41</sup> | 77,090 females from Nurses' Health Study, mean age range 47.7-51.7 years across quintiles of legume intake. USA. | Legume quintiles as weekly servings ranging <0.5 to >2. | Self-reported cholecystectomy | = | Acceptable |
| Case-control studies |  |  |  |  |  |
| Jayanthi et al.<br>(2005)<sup>57</sup> | 346 cases and 346 sex and age-matched controls, 51.4% females, mean age cases: 51 years, controls: 50.3 years. India. | Consumption frequency of Tamarind bean paste | Gallstone | > | Unacceptable |
| Parveen et al.<br>(2019)<sup>59</sup> | 100 cases and 100 sex and age-matched controls, 74% female cases, 44% female controls, mean age not presented. Pakistan. | Weekly legume intake | Gallstone | ≥ | Unacceptable |
| Pastides et al. | 84 gallstone cases, 171 controls, 100% females, mean age | Monthly legume intake | Gallstone | = | Unacceptable |
| (1990)<sup>60</sup> | cases: 54.8 years, controls: 54.3 years. Greece. | frequency |  |  |  |
| Rasheed et al.<br>(2020)<sup>58</sup> | 157 gallstone cases, 175 healthy controls, 100% females, mean age not presented. Saudi Arabia. | Legume intake | Gallstone | > | Unacceptable |
| Thijs et al.<br>(1990)<sup>25</sup> | 204 gallstone cases and 615 controls, 67.7% female cases, 69.4% female controls, mean age not presented. The Netherlands. | Legume intake<br>frequency | Gallstone | < | Unacceptable |
| Cross-sectional studies |  |  |  |  |  |
| Tseng et al.<br>(2000)<sup>61</sup> | 4641 Mexican American participants from NHANES III, 49.7% females, mean age women: 37.5 years, men: 35.8 years. USA. | Weekly legume intake<br>frequency | Gallstone and<br>cholecystectomy | ≥ | Low |
| Unisa et al.<br>(2010)<sup>62</sup> | 6548 participants, symptomatic patients, 59.9% females, mean age not presented. India. | Chickpea intake<br>(Yes/No) | GBD<sup>d</sup> and<br>gallstone | ≥ | Very low |

<sup>a</sup>Results are presented as “<” corresponding to an inverse association, “=” corresponding to no association, “≥” corresponding to a positive or no association, and “>” corresponding to a positive association. <sup>b</sup>Quality refers to the methodological quality assessment of the study. <sup>c</sup>E3N, Etude Épidémiologique auprès de femmes de la Mutuelle Générale de l'Education Nationale. <sup>d</sup>GBD, gallbladder disease

### Legumes and risk of NAFLD

One study investigated the longitudinal association of replacing animal-based proteins with legumes in a large sample of individuals from Tianjin Chronic Low-grade Systemic Inflammation and Health Cohort (TCLSIH) and Guangzhou Nutrition and Health Study (GNHS), two large national cohorts in China. ^40^ Substituting poultry for legumes was associated with a lower HR of NAFLD (HR: 0.35, 95% CI: 0.18; 0.69, and HR: 0.92, 95% CI: 0.86, 1.00 in GNHS and TCLSIH respectively). ^40^ Substituting one serving of processed meat with one serving of legumes was associated with a lower risk of NAFLD in TCLSIH (HR: 0.86, 95% CI: 0.74; 1.00), while the GNHS did not differentiate between red and processed meat and found a higher risk of NAFLD when replacing one serving of red meat with one serving of legumes (1.16, 95% CI: 0.99; 1.37). ^40^ The methodological quality was evaluated as high (Appendix V).

Five case-control studies found heterogeneous results. Bahrami et al. ^43^ investigated the association between increasing legume consumption by 1 serving/week and NAFLD among 196 Iranian cases and 803 controls. The association for beans, lentils, and total legumes were each assessed separately and showed a significantly lower odds ratio (OR) for NAFLD when increasing the consumption with 1 serving/week across any legume category. Giraldi et al. ^46^ investigated the association between the Mediterranean dietary pattern and NAFLD among 371 Italian individuals with NAFLD and 444 controls. In sub-analyses, the authors found that the highest compared to lowest legume consumption was associated with lower OR for NAFLD (OR: 0.62, 95%CI: 0.38; 0.99). ^46^ Han et al. ^47^ assessed the association between daily bean intake and NAFLD in 169 cases and 179 controls. The odds for developing NAFLD did not differ between the lowest and highest tertile of bean intake (OR for men: 1.50, 95% CI: 0.59; 3.84, OR for women: 1.13, 95% CI: 0.44; 2.89). ^47^ Hao et al. ^48^ investigated the association between the consumption frequency of beans and NAFLD in individuals with different genotypes for methylenetetrahydrofolate reductase (MTHFR). They found lower OR for NAFLD with higher consumption frequency of beans. However, the study only included information on weekly frequency (OR: 0.701, 95% CI: 0.576; 0.853). ^48^ There was some evidence of differing associations by MTHFR-genotype (OR for CC genotype: 0.807, 95% CI: 0.541; 1.204, OR for CT+TT genotype: 0.670, 95% CI: 0.535; 0.839). ^48^ Tutunchi et al. ^49^ found little evidence of an association between legume consumption and NAFLD (OR: 0.74, 95% CI: 0.61; 1.67) among 105 Iranian cases and 105 controls. The quality assessment of the case-control studies ranged from high to poor (Appendix V).

The nine cross-sectional studies investigating the association between legume consumption and NAFLD generally found inverse associations, often not statistically significant. Baratta et al. ^44^ found little evidence of an association between higher legume consumption and NAFLD among 584 Italian participants (OR: 0.675, 95% CI: 0.370; 1.230). Bullón-Vela et al. ^50^ included 328 Spanish individuals with MetS and found that the highest compared to lowest tertile of legume consumption (g/day) was associated with a lower relative risk ratio (RRR) for high liver fat content (RRR: 0.48, 95%CI: 0.24; 0.97). Chan et al. ^45^ investigated the association between soy product consumption and NAFLD among 797 Chinese individuals and found little evidence of an association (OR: 0.86, 95% CI: 0.52; 1.42). Chiu et al. ^42^ investigated substituting one serving of meat with one serving of soy in 3279 Taiwanese individuals and found no association with NAFLD (OR: 0.95, 0.88; 1.02). Mirizzi et al. ^51^ included baseline information on 136 individuals from a 6-arm RCT study and observed evidence of an association between chickpea consumption and NAFLD severity (OR: 0.57, 95%CI: 0.34; 0.97), but not for soy milk and dried peas. Vijay et al. ^52^ observed some evidence of an association between consumption of dried pulses and legumes and NAFLD among Indian individuals (OR: 0.43, 95% CI: 0.21; 0.61). Watzinger et al. ^53^ found little evidence of an association comparing the lowest and highest quintiles of legume consumption and NAFLD among 136 German individuals recruited for the HELENA trial (OR: 1.78, 95% CI: 0.57; 5.55). In a Japanese sample of 431 participants, Yabe et al. ^54^ observed little evidence of an association between consumption of soybeans or soybean products ≥ 4 times/week compared to once weekly and NAFLD (OR 0.57, 95% CI: 0.27;1.19). Zhang et al. ^39^ stratified their analyses according to sex, age, and BMI, and found some evidence of an association between soy food intake ≥4 times/week compared to <1 time/week and NAFLD for age < 50 years (OR: 0.70, 95% CI: 0.59; 0.84), BMI ≥ 24 kg/m2 (OR: 0.76, 95% CI: 0.65; 0.89), and men (OR: 0.73, 95% CI: 0.61; 0.87). The cross-sectional studies were evaluated to be of varying methodological quality, ranging from poor to high, primarily due to missing information on selection procedures. Some studies validated their measurement methods, while others did not, but considerations regarding study design or adjustment levels to account for confounding factors were often poorly described or inadequately implemented (Appendix V).

Three cross-sectional and one case-control study were summarized in a systematic review investigating various food groups and their relation to NAFLD. ^3^ These studies were also included in this current scoping review. ^42–45^ The pooled analysis of the cross-sectional studies observed little evidence of an association between legume consumption and NAFLD (OR: 0.943, 95% CI: 0.877; 1.014). ^3^ The confidence in the results from the review was evaluated as moderate (Appendix V).

One narrative review proposed that the mechanisms underlying the potential association between legumes and NAFLD were due to alterations in glucose and lipid metabolism. ^55^ This was based on animal studies indicating that a globulin, beta-conglycinin from soy, downregulates the expression of PPARγ-2 in animal models, which has been associated with obesity and storage of lipids in the liver. ^65^ The review presented findings from Bahrami et al. ^43^ and an RCT of 42 premenopausal women with central obesity who experienced decreased levels of the liver health biomarkers aspartate aminotransferase (AST) and alanine aminotransferase (ALT) when consuming a hypocaloric diet with legumes for six weeks compared to a hypocaloric diet without legumes. ^55^

### Legumes and risk of GBD

Barré et al. ^56^ investigated the risk of gallbladder removal in the French E3N cohort and found evidence of lower risk (HR: 0.73, 95% CI: 0.65; 0.82) among those consuming more than 27.8 g/day compared to 0 g/day. Another prospective cohort using data from the Nurses’ Health Study across 1,060,033 person-years did not find clear evidence of an association between legumes and gallbladder removal (HR: 1.02, 95%CI: 0.94; 1.12). ^41^ The studies were evaluated as having high and moderate study quality, respectively (Appendix V).

Five case-control studies investigated the association between legume consumption and GBD and observed mixed directions of associations. Jayanthi et al. ^57^ investigated the odds of gallstone formation based on the consumption frequency of tamarind bean paste among 346 cases and 346 controls in India. They observed evidence of a positive association between consuming bean paste ≥3 times weekly compared to <3 times weekly (OR: 1.79, 95% CI: 1.09; 2.93). Parveen et al. ^59^ included 100 cases and 100 controls from Pakistan and also observed a positive association between consuming peas once weekly and the development of gallstones (OR: 3.37, 95% CI: 1.20; 9.60). Pastides et al. ^60^ did not observe evidence of an association in Greek women between legume consumption and gallstones in a linear trend analysis (chi-squared: 2.97, p=0.085) among 84 cases and 171 controls, while Rasheed et al. ^58^ found a significantly higher intake of legumes among 157 gallstone cases compared to 175 healthy controls (consumption among cases 43.3 %, controls 33.3 %, p<0.05) in females from Saudi Arabia. Thijs et al. ^25^ observed an inverse association between legume consumption frequency and gallstones in a case-control study of 204 cases and 615 controls from the Netherlands. Consuming legumes eight or more times monthly compared to less than once monthly was associated with significantly lower odds of gallstones (OR: 0.4, 95%CI: 0.23; 0.87). ^25^ The large variation in results may be caused by the methodological quality, which was rated as poor for all studies (Appendix V).

Two cross-sectional studies investigated the association between legume consumption and GBD. Tseng et al. ^61^ investigated the prevalence OR (POR) of GBD among 4641 Mexican Americans from NHANES III by comparing the consumption frequency of legumes and observed only weak evidence of an association among those consuming beans every day compared to less than once weekly (POR: 0.59, 95% CI: 0.24; 1.44). However, when stratifying the sample on gender and only including individuals who were unaware of having prevalent gallstones, women consuming legumes daily compared to less than once weekly had a higher POR for gallstones (POR: 1.58 (1.02; 2.46). Unisa et al. ^62^ observed evidence of a positive association between chickpea consumption and gallstone prevalence among men (OR: 2.55, 95% CI: 1.56; 4.15) but only weak evidence among women (OR: 1.35, 95% CI: 0.98; 1.86) in a sample of 6548 Indian participants. The methodology of the studies was rated as low and very low, respectively (Appendix V).

Three narrative reviews^21,63,64^ suggested that consuming legumes may be linked to a higher occurrence of gallstones since the prevalence of gallstones is very high among populations who consume legumes such as beans as a dietary staple. The prevalence of gallstone and GBD can partly be attributed to cholesterol supersaturation of the bile caused by high legume intakes. ^63,64^ Being overweight and obese significantly increases the risk of gallstone formation and 75% of all gallstones in Western countries are related to metabolic abnormalities such as type 2 diabetes, MetS, and NAFLD. ^63^ Gaby et al. ^21^ further described that food allergies may play a role in inhibiting gallbladder emptying and thus modify the association between certain foods and GBD, which may pose an additional risk in individuals with food allergies.

## Discussion

The literature on the association between legume consumption and NAFLD indicated some evidence of a lower risk of NAFLD with high legume consumption. Ten articles reported an inverse association, while seven articles reported no clear association. However, the methodological quality of the included studies was generally low or moderate and high for only three. This warrants further research with more transparent reporting in order to draw conclusions on the association between legumes and NAFLD risk with greater certainty.

For GBD, the literature was inconclusive of the direction of associations. Two articles found an inverse association, two found no association, and five found a positive association between legume consumption and GBD. The methodological quality of the included studies was generally low, with only one study evaluated as of high quality. The discrepant results and generally poor study quality contribute to the lack of clear evidence of an association between legume consumption and GBD.

The large variation in results may be caused by bias introduced by poor methodological quality of the studies lacking information on case ascertainment, validation of diet information, and small sample sizes, but may also result from the inclusion of populations with different food cultures. Previous research has indicated that plant-based meat alternatives are understudied in relation to health. ^66,67^ No studies in this review investigated the association between plant-based meat alternatives and NAFLD or GBD, and this association was not evaluated.

## Dietary assessment

Short-term dietary assessments may not be fit for estimating habitual dietary intake, despite offering detailed information. ^68,69^ Particularly for periodically consumed foods, a food frequency questionnaire (FFQ) may be a more efficient for capturing habitual dietary intakes. ^69,70^ However, habitual diet can be equally well estimated when using three or more short-term dietary assessments. ^71^ Measurement errors are not uncommon and the diet assessment tool should therefore be validated as an appropriate tool to measure dietary exposure in the population of interest. ^69,70,72^ The validity of an FFQ is often evaluated against another diet assessment method with 24-hour recalls or food diaries being most common due to their high detail level for quantification of intakes. ^68–71^ Biomarker validation has also emerged in recent years to assess nutrient intakes. ^70,72,73^ The strength in validating different diet assessment methods against each other, and potentially also against biomarkers, is the differences in potential measurement errors between assessment methods. ^69–71,73^

### Studies on NAFLD

Most studies on NAFLD used FFQs to assess dietary intakes. ^39,40,42–46,48,50–54^ Several studies evaluated reproducibility between two FFQs filled in at least one year apart. Furthermore, the FFQs were most often validated against other dietary assessment methods or biomarker assessments of nutrient intake. Some studies used validated, but poorly described or designed, FFQs while others again did not use validated methods to assess dietary intakes.

The prospective follow-up study and one case-control study used validated FFQs to assess dietary intakes, ^40,43^ while two case-control studies used non-validated FFQs. ^46,48^ Two case-control studies used short-term dietary assessments without validation, ^47,49^ offering only snapshots of dietary intakes potentially inadequate for investigating long-term diet-disease associations. ^68^ There was no clear pattern between validated dietary assessments and the association between legume consumption and NAFLD among the retrospective or prospective studies. The two studies with validated FFQs^40,43^ found differing results like the studies using non-validated FFQs, which also reported inverse or no association. ^46,48^ The studies using short-term diet assessment found no association. ^47,49^ Six cross-sectional studies used validated FFQs, ^39,42,44,50,51,53^ and three used non-validated FFQs. ^45,52,54^ The magnitude and direction of associations reported in studies with validated FFQs were not uniform and differing results were found. Those employing non-validated FFQs all found no association between legume consumption and NAFLD. This could indicate that the non-validated FFQs employed in the cross-sectional settings may not adequately estimate habitual dietary intake and thus cause non-differential misclassification biasing the estimate towards the null.

There was no clear pattern between the validation of dietary assessment and the association between legume consumption and NAFLD. The direction and magnitude of associations varied across study designs and the validity of diet assessments. There was some indication that using non-validated short-term dietary assessments and cross-sectional studies using non-validated FFQs resulted in non-significant associations. The lack of clear and validated diet assessments raises doubts on their accuracy, raising concerns that they may not accurately reflect habitual legume intake or the true association between legume consumption and NAFLD. Furthermore, retrospective case-control studies and cross-sectional studies can be particularly susceptible to differential misclassification as exposure and outcome assessments may affect each other. ^74^ Despite some studies not validating their diet assessment methods, many studies using valid FFQs found non-significant associations between legume consumption and NAFLD. ^40,42,44,51,53^ The studies employing non-validated exposure assessments did, however, mostly find inverse associations between legume consumption and NAFLD, which suggests that methodological differences in exposure assessment may in part drive the divergent findings.

### Studies on GBD

The two prospective cohorts investigating GBD used FFQs validated against a short dietary assessment method^41,56^ but found different magnitudes of associations indicating an inverse association or no association between legume consumption and GBD. Of the five case-control studies, four used non-validated dietary assessment methods, ^25,57,59,60^ while one failed to include information on the validation. ^58^ Two studies used questionnaires with one collecting information on diet together with other factors for GBD, ^58^ while the other did not include details on the questionnaire apart from the description of one question regarding the consumption frequency of beans and legumes over the past month. ^25^ Three studies assessed dietary intake through interviews, ^57,59,60^ of which two studies also assessed other factors related to GBD during the diet assessment interviews. ^59,60^ Despite the heightened precision in diet assessment through trained personnel conducting the interviews, the interviews as a tool were not validated. This also raises concerns about the ability to assess intakes of all legume foods or quantities consumed, underscoring the necessity for a comprehensive food intake evaluation to ensure a valid measure of habitual intake. These issues could potentially explain the heterogeneity in results due to different levels of misclassification. This renders some uncertainty as to whether the estimated association reflects the true association between legume consumption and GBD. The two cross-sectional studies did not use validated dietary assessment methods or failed to describe the validation. ^61,62^ One study adapted a previously validated FFQ to its study population, ^61^ while the other study did not account for the validity of their interview-based approach. ^62^

The poor validation of dietary assessment or lack thereof across the different study designs may have introduced information bias, rendering uncertainty about the estimated associations’ ability to reflect the true association between legume consumption and GBD. However, the studies using valid diet assessments also found different directions and magnitudes of association, indicating that the dietary assessment validation might not be a main driver for the diverging results across studies.

## Outcome assessment

Liver biopsies are the most accurate tool for diagnosing NAFLD, but also invasive and painful. Less invasive methods includes liver imaging such as ultrasound or magnetic resonance imaging (MRI), which have been found valid for diagnosing NAFLD with high specificity and precision when images are interpreted by a gastroenterological or hepatological expert. ^75–78^ The standard criteria for the non-invasive diagnosis of NAFLD require an indication of a “bright liver pattern” with a hepatic fat content of ≥5% in the absence of excessive alcohol consumption. ^75–78^

The most common assessment of gallstones is ultrasound examination, which has been validated to have high specificity and sensitivity for detecting gallstones when conducted by a trained specialist. ^79^ Since the most common treatment of symptomatic gallstones is surgical removal, cholecystectomy, the presence of gallstones will be diagnosed directly during the surgery for most cases. ^79^

### Assessment of NAFLD

Several of the studies used validated NAFLD assessment methods. Seven studies used the standard “bright liver pattern” criteria for diagnosing NAFLD with ultrasound or MRI imaging evaluated by one or more trained hepatologists or gastroenterologists. ^43–46,48,49,53,75–77^ Seven studies defined NAFLD based on fat deposition in the liver assessed through echogenicity or elastography conducted by experienced technicians. ^39,40,42,43,51,52,54,75–78^ One study employing ultrasound imaging did not clarify the criteria for NAFLD diagnosis, ^47^ and another used a non-imaging technique to diagnose NAFLD with the hepatic steatosis index. ^50^ This method incorporates ALT:AST ratio, BMI, diabetes status, and female sex and has been validated against liver biopsies for NAFLD diagnosis. ^50,80^ Two studies supported their imaging technique by using composite diagnosis methods incorporating levels of ALT and AST. ^43,54^

All studies, except one, reported validated outcome measures to diagnose NAFLD and differences in results across studies can hardly be attributed to the outcome assessment methods.

### Assessment of GBD

The included studies in GBD used three outcome assessment methods: surgery, imaging, and self-reported outcomes. The two prospective cohorts used self-reported cholecystectomy as outcomes. ^41,56^ Both studies validated the outcome against medical records with high reliability and validity. Furthermore, self-reported cholecystectomy increases accuracy in reporting compared to self-reported gallstones, as gallstones may be non-symptomatic. Three case-control studies used validated and non-invasive imaging to diagnose gallbladder diseases, ^25,58,60^ while two studies failed to clarify their diagnostic criteria. ^57,59^ This did not seem to impact the observed association between legume consumption and GBD, as the direction and magnitude varied across all studies independent of valid or non-valid outcome assessments. The two cross-sectional studies used valid outcome assessments with ultrasound imaging and expert radiologists interpreting the images. ^61,62^ As the studies found opposite directions of associations between legume consumption and GBD, the differences cannot be ascribed to the outcome assessment methods.

Poor description of diagnostic criteria or lack of validation raises uncertainty about diagnoses, which could potentially cause misclassification and biased estimates. Despite some studies poorly describing outcome assessment methods, there was no clear pattern between the direction and magnitude of associations between legume consumption and GBD across the validity of outcome assessments. Furthermore, most studies had ultrasound experts or senior radiologists assessing the images to diagnose GBD providing some certainty in the outcome assessments. Erroneously defined outcomes do not appear to be the major driver of the different study results.

## Adjustment strategies

### Studies on NAFLD

The association between diet and NAFLD is prone to confounding from age, sex, alcohol consumption, physical activity, obesity or overweight, and metabolic factors such as dyslipidemia, MetS, and type 2 diabetes. ^39,40,42,45,47,48,52,54^ All studies accounted for age, while three studies did not account for sex. ^47,48,50^ Several studies failed to adjust or account for important confounding factors like alcohol consumption, ^43,48^ ^53^ physical activity, ^44,45,48,51,52^ ^53^ obesity or overweight, ^47,48,51,54^ and metabolic factors. ^48,49,51,52,54^ Furthermore, seven studies did not adjust for other foods consumed, ^45–49,52^ ^53^ and only six studies accounted for SES. ^39,40,42,47,49,52^ The context in which a person lives his or her general life has in various research been shown to be important in relation to diet and health. Living alone has been associated with poorer diet quality and poorer health outcomes than cohabiting with others. ^81,82^ It could be assumed individuals living alone may also be at greater risk of developing NAFLD due to poorer dietary habits and overall poorer health. Therefore, not adjusting for cohabitation might bias the estimated association between diet and NAFLD toward the null.

Several of the studies failed to account for important confounders of the association which may bias estimates towards and away from the null. This raises uncertainty that the estimated associations reflect the true associations between legume consumption and NAFLD due to residual confounding, which could partly explain the difference in results, with the majority being non-significant potentially due to residual confounding biasing the estimates towards the null.

### Studies on GBD

Important confounding factors related to the association between diet and GBD are age, sex, parity, hormonal drug use for women, pregnancy, physical activity, smoking, and obesity, overweight, or rapid weight loss. ^15,41,56,58–63^ Several studies failed to account for many of these confounding factors. Two studies did not mention adjustment levels, and both of these found that higher legume consumption was associated with greater risk of GBD. ^57,58^ Four studies accounted for age and sex through design or adjustments but failed to otherwise include or describe the inclusion of other important covariates. ^25,59,60,62^ Despite one of the studies trying to increase the comparability of controls and cases through different matching strategies for acute and non-acute cases, ^25^ all studies had substantial risk of residual confounding. This might potentially explain the opposite directions of associations found, with one study finding a positive association between legume consumption and GBD, ^62^ one finding an inverse association, ^25^ and two finding no association. ^59,60^ Some studies accounted for a multitude of potential confounding factors including sex, age, dietary habits, weight loss, smoking, overweight or obesity, hormonal drug use for women, parity, pregnancy, etc. ^41,56,61^ Despite the various adjustment levels, important factors like physical activity, SES, and cohabitation were not included. Furthermore, the obtained results were of different direction and magnitude implying that the multiple-adjusted analyses did not obtain more uniform associations than the poorly adjusted analyses. One of the studies failed to present the results of the multivariable-adjusted analysis, ^61^ raising concerns about residual confounding and bias of the estimated association.

The large variation in adjustment for confounders together with differences in outcome definitions render the certainty of the estimated associations unclear due to the substantial risk of residual confounding. All studies failed to include important confounding factors such as physical activity, SES, parity, cohabitation, and rapid weight loss. The differences in adjustment levels and statistical analyses can most likely explain the diverging results across studies.

## Population differences

This review encompasses a broad range of population samples with undoubtedly different food cultures. Diet is shaped by local culture and varies between ethnicities, cultural groups, and genders, and may also vary because of personal preference and food availability. ^83^ Furthermore, SES and cohabitation have been shown to impact an individual’s dietary habits and health. ^81,82,84^

### Studies on NAFLD

For the association between legume consumption and NAFLD there appears to be some distinction between food cultures. The Asian cuisine and food culture traditionally includes more legumes and an overall healthier diet compared to Western diets. ^85,86^ However, the studies with Asian populations^39,40,42,45,47,48,52,54^ did not find clear associations between legumes and NAFLD. Results were of somewhat diverging directions, and magnitudes indicated that the cultural meal setting, and foods consumed in combination with legumes may be important for the risk of developing NAFLD. There appear to be promising hepatobiliary health benefits for the studies including Middle Eastern and Mediterranean populations. Legumes are a staple food in many Middle Eastern countries, and both studies with Middle Eastern populations found inverse associations between legume consumption and NAFLD. ^43,49^ Furthermore, the foods typically consumed together with legumes in the Mediterranean areas such as fish, plant-based fats, and nuts may provide an optimal combination of food matrices and nutrients to lower the risk of NAFLD. All studies with Mediterranean populations found inverse associations between legume consumption and NAFLD. ^44,46,50,51^ Meanwhile, foods typically consumed in combination with legumes in Central Europe may to a greater extent include animal-based fats from butter, lard, or dairy, and coarse vegetables like kale, cabbage, and onion, which could explain the positive association between legume consumption and NAFLD in the study including a Central European population. ^53^

### Studies on GBD

The combination of foods consumed, which is highly dependent on food culture and tradition and varies across geographical locations, may significantly impact the association between legume consumption and GBD. The studies including populations with tradition of consuming pulses such as Asian^57,59,62^ and Middle Eastern^58^ populations indicated that legume consumption was associated with a greater risk of GBD. However, one study included a population of Mexican Americans, ^61^ who also have tradition of consuming beans, ^87^ but did not find any association between legume consumption and GBD. This study sampled their participants in the USA with an oversampling of Mexican Americans. Research has indicated that immigration often results in altered dietary behavior often through the inclusion of foods with high fat and sugar contents and low nutrient density while excluding nutrient-dense foods such as vegetables, fruits, and legumes. ^88^ Immigration to the USA might have altered participants’ dietary habits to resemble Western dietary patterns with a lower consumption of legumes and pulses compared to native Mexican citizens still living in Mexico. The results of the studies including Western Populations^25,41,56,60^ indicated that the Westernized population might not be at increased risk of developing GBD when consuming legumes, and there might even be an inverse association. Overall, there appears to be a tendency that populations with a generally greater tradition for consuming legumes and pulses are at greater risk of developing GBD. Whether the legumes drive this association alone, or it is the combination of foods in which legumes are consumed is unclear.

## Underlying mechanisms

Legumes are well established as a health-promoting dietary factor due to their high contents of antioxidants, polyphenols, dietary fibers, and bioactive peptides. ^8,13,55^ Several hepatobiliary mechanisms have been proposed to be affected by legumes, including digestion, lipid metabolism, and cholesterol synthesis. Dietary patterns with a high content of legumes have been associated with a lower risk of NAFLD. ^11,12,18,55^

Consumption of legumes is linked to lower serum lipids by affecting digestion and bile acid metabolism. ^14–17^ Dietary fibers, abundant in legumes, bind cholesterol in the intestine, aiding its removal from the body. Soluble fibers slow down gastric emptying and nutrient absorption, reducing the uptake of fats like TG and free fatty acids. ^55,89,90^ Fibers from legumes also bind bile acids, reducing their reabsorption into the liver and prompting increased bile acid production, which in turn lowers hepatic cholesterol levels and serum cholesterol through negative feedback. ^14,89,90^ Additionally, bioactive compounds in legumes can lower cholesterol through lipid metabolism and lipid reabsorption prevention. ^89^ Simultaneously, resistant starches from legumes can be fermented by colon bacteria to produce beneficial short-chain fatty acids like propionate and butyrate. These fatty acids can promote a favorable environment for beneficial colon bacteria, which may affect the lipid metabolism through altered fat oxidation, lipogenesis, and lipolysis. ^55,90^

Fat is primarily stored in the liver as TG and individuals with elevated serum levels typically have an overproduction in the liver combined with impaired TG removal from the blood. ^89^ Triglycerides are transported in very low-density lipoproteins (VLDL) and excreted from the liver into the bloodstream through the intestinal tract. ^89^ Increased bile acid secretion and inhibited reabsorption in the intestines can bind TG, facilitating its excretion in feces. ^90,91^ These mechanisms lower the availability of circulating lipids that can be stored in the liver tissue and can thus minimize the risk of NAFLD.

Consuming legumes as a diet staple is associated with a high prevalence of gallstones. ^15,20,21,64^ However, proposed triggers of GBD also include other dietary factors and metabolic issues, and even some genetic factors have been found to promote the development and progression of gallstones. ^15,19,63,92^ NAFLD progression affects bile duct and gallbladder function by expelling surplus lipid fractions via the gallbladder. ^15,19,63^ Gallstone formation is a multifaceted process, with cholesterol supersaturation of bile being a key factor. ^15,20,21,61,64^ Bile salts combine with phospholipids and cholesterol to boost cholesterol solubility, but an imbalance in lipids and bile salts can lead to supersaturation and crystal formation. Elevated cholesterol levels relative to bile salts and phospholipids lead to cholesterol supersaturation in bile, fostering crystal precipitation. Increased cholesterol secretion into bile and reduced bile salt secretion are two metabolic processes contributing to cholesterol supersaturation. ^15,20,21,64^ Furthermore, a large proportion of gallstones in Western countries has been related to metabolic abnormalities linked to overweight and obesity, including NAFLD. ^63^

### Strength and limitations

This scoping review has several strengths including the rigorous literature search strategy. To our knowledge, the scoping review framework allowed for the inclusion of all literature of relevance to investigate the association between legume consumption and hepatobiliary health. The review process was extensive including the old nomenclature for NAFLD and the new nomenclature for MASLD, while evaluating all available data sources not only limited to those with an evidence level. The methodological assessment of evidence-level articles provided a tool to critically assess the quality of available research when interpreting the estimated associations.

A scarcity of research in this area led us to broaden the scope of this study, thus including studies focusing on various aspects of hepatobiliary disorders. The studies were diverse and heterogeneous, using a wide variety of methods and approaches. The evidence was thus challenging to synthesize into a clear demonstration of an association between legume consumption and NAFLD or GBD.

## Conclusions

Twentynine articles investigating the association between consumption of legumes were included in this review. Seventeen of these investigated NAFLD-outcomes (1 cohort study, 5 case-control studies, 9 cross-sectional studies, 2 reviews) and 12 investigated GBD-outcomes (2 cohort studies, 5 case-control studies, 2 cross-sectional studies, 3 reviews).

Despite some literature indicating a protective association between legumes and NAFLD, the results were heterogeneous, and a clear association can therefore not be established. The methodology of the included studies has substantial differences in relation to the assessment of exposures, and the statistical analyses may have led to diverging results due to different covariate adjustment strategies. The overall confidence in the evidence varied and more clearly designed and reported research on the topic is warranted.

For gallbladder diseases, the literature was diverging with different magnitudes and directions of associations rendering a clear conclusion impossible as legume consumption was reported to be inversely, positively, or not associated with gallbladder diseases.

Methodological differences between studies warrant more clearly designed and reported research. The potential for legume consumption to increase gallstone risk should, however, be considered in light of the recognized health benefits associated with the consumption of legumes.

Due to the heterogeneity of the results of this review, any proposed underlying mechanisms between legume consumption and the development of NAFLD or GBD cannot be confirmed. Future research investigating associations and underlying mechanisms could potentially fill a substantial knowledge gap and provide important insights into the diet-disease interactions and underlying pathways.

## Acknowledgments

Author contributions

FL and CCD were responsible for designing the research question. FL designed the search strategy with guidance from CCD. FL and CCD drafted the protocol, data extraction template, and results template, VLM, LR, and NB critically reviewed the protocol and relevant templates. FL conducted the preliminary search. FL, VLM, CFJ, LR, and NB conducted the literature screening and full-text review. Data extraction and bias assessment were done by FL, VLM, CFJ, LR, NB, and CCD. Tabular descriptions of results were done by FL. All authors critically revised the manuscript and take full responsibility for the content of this scoping review.

## Data availability

A complete list of all literature identified with the search strategy presented in Appendix I, after the automatic exclusion of duplicates in Covidence, can be requested from the corresponding author.

## Supporting Information

Appendix I: Search strategy

Appendix II: Search strategy for unpublished and grey literature

Appendix III: Studies ineligible following full-text review

Appendix IV: Data extraction instrument

Appendix V: Methodological quality assessment of included studies

Appendix VI: Characteristics of included studies

Appendix VII: PRISMA-ScR Checklist

