## Appendices for "Consumption of legumes and risk of hepatobiliary diseases among humans aged 2+ years: a scoping review"

Christina C. Dahm

### Appendix I: Search strategy

PubMed (National Center for Biotechnology Information, MD USA)
Search conducted on July 24th, 2023.

| Search | Query | Records retrieved |
|---|---|---|
| #1 | ((((((((((((((((((((fabaceae[MeSH Terms]) OR (lens plant[MeSH Terms])) OR (peas[MeSH Terms])) OR (legume*)) OR ("dietary pulse*")) OR ("legume pod*")) OR ("pod*, legume*")) OR (lentil*)) OR (bean*)) OR (pea*)) OR (fabaceae)) OR ("vigna"[All Fields])) OR ("phaseolus vulgaris"[All Fields])) OR ("phaseolus mungo"[All Fields])) OR ("black gram"[All Fields])) OR ("red gram"[All Fields])) OR ("chickpea"[All Fields])) OR ("bengal gram"[All Fields])) OR ("phaseolus"[All Fields])) OR ("soybean"[All Fields])) OR ("glycine max"[All Fields]) | 173,087 |
| #2 | (((((((((((((((((((((((((cholestasis[MeSH Terms]) OR (cholecystectomy[MeSH Terms])) OR (cholecystitis[MeSH Terms])) OR (acalculous cholecystitis[MeSH Terms])) OR (choledocholithiasis[MeSH Terms])) OR (cholelithiasis[MeSH Terms])) OR (liver cirrhosis, biliary[MeSH Terms])) OR (biliary tract[MeSH Terms])) OR (gallstone*[All Fields])) OR ("gallbladder disease*"[All Fields])) OR ("bile duct gallstone"[All Fields])) OR ("common bile duct gallstone"[All Fields])) OR ("gallbladder inflammation"[All Fields])) OR ("gallbladder inflammatory"[All Fields])) OR ("acalculous cholecystitis"[All Fields])) OR ("acalculous gangrenous cholecystitis"[All Fields])) OR (cholelithias*[All Fields])) OR ("choledocholithiasis"[All Fields])) OR ("choledocholithias*"[All Fields])) OR ("cholecystectomy"[All Fields])) OR ("cholecystectomies"[All Fields])) OR ("gallbladder removal"[All Fields])) OR ("removal of gallbladder"[All Fields])) OR ("biliary colic"[All Fields]) ) OR ("biliary calculi"[All Fields])) OR ("biliary calculosis"[All Fields]) | 165,909 |
| #3 | (((((((("Non-alcoholic Fatty Liver Disease"[Mesh]) OR (gastroenterology[MeSH Terms])) OR ("non alcoholic fatty liver"[All Fields])) OR ("non alcoholic fatty liver disease"[All Fields])) OR ("non alcoholic steatotic hepatitis"[All Fields])) OR ("non alcoholic steotohepatitis"[All Fields])) OR ("nafld"[All Fields])) OR ("nash"[All Fields])) OR ("liver"[MeSH Terms]) | 524,308 |
| #4 | #2 OR #3 | 661,089 |
| #5 | #1 AND #4 | 4047 |
| | Limited to dates from 1000/1/1 to 2023/6/30 | |

PubMed (National Center for Biotechnology Information, MD USA)
Search conducted on July 24th, 2023.

| Search | Query | Records retrieved |
|---|---|---|
| #1 | (((((((((((((((((((fabaceae[MeSH Terms]) OR (lens plant[MeSH Terms])) OR (peas[MeSH Terms])) OR (legume*)) OR ("dietary pulse*")) OR ("legume pod*")) OR ("pod*, legume*")) OR (lentil*)) OR (bean*)) OR (pea*)) OR (fabaceae)) OR ("vigna"[All Fields])) OR ("phaseolus vulgaris"[All Fields])) OR ("phaseolus mungo"[All Fields])) OR ("black gram"[All Fields])) OR ("red gram"[All Fields])) OR ("chickpea"[All Fields])) OR ("bengal gram"[All Fields])) OR ("phaseolus"[All Fields])) OR ("soybean"[All Fields])) OR ("glycine max"[All Fields]) | 173,087 |
| #2 | ((("metabolic dysfunction-associated fatty liver disease") OR ("metabolic dysfunction-associated steatohepatitis")) OR (MAFLD)) OR (MASH) | 4032 |
| #3 | #1 AND #2 | 160 |
| | Limited to dates from 1000/1/1 to 2023/6/30 | |

Embase (Elsevier B.V., Amsterdam, Netherlands
Search conducted on July 24th, 2023.

| Search | Query | Records retrieved |
|---|---|---|
| #1 | 'legume'/exp OR 'chickpea' OR 'lentil' OR 'grain legume' OR 'bean' OR 'fabaceae' OR 'pea' OR 'legume pod' | 132,115 |
| #2 | 'hepatology'/exp OR 'liver'/exp OR 'liver disease'/exp OR 'gallbladder'/exp OR 'acalculous gallbladder inflammation' OR 'gallstone'/exp OR 'cholelithiasis'/exp OR 'bile duct'/exp OR 'bile duct disease'/exp OR 'common bile duct gallstones' OR 'acute cholecystitis'/exp OR 'cholecystectomy'/exp OR 'fatty liver'/exp OR 'gallbladder disease' OR cholecystitis OR 'nonalcoholic fatty liver' OR 'removal of gallbladder' | 1,968,923 |
| #3 | #1 AND #2 | 3623 |
| | Limited to dates from 1000/1/1 to 2023/6/30 | |

Embase (Elsevier B.V., Amsterdam, Netherlands
Search conducted on July 24th, 2023.

| Search | Query | Records retrieved |
|---|---|---|

| | | |
|---|---|---|
| #1 | 'legume'/exp OR 'chickpea' OR 'lentil' OR 'grain legume' OR 'bean' OR 'fabaceae' OR 'pea' OR 'legume pod' | 132,115 |
| #2 | 'metabolic fatty liver'/exp OR 'metabolic fatty liver' OR 'metabolic dysfunction associated steatotic liver disease'/exp OR 'metabolic dysfunction associated steatotic liver disease' OR 'metabolic dysfunction associated steatohepatitis'/exp OR 'metabolic dysfunction associated steatohepatitis' | 1415 |
| #3 | #1 AND #2 | 3 |
| | Limited to dates from 1000/1/1 to 2023/6/30 | |

CINAHL complete (EBSCO Information Services, MA, USA)
Search conducted on July 24th, 2023.

| Search | Query | Records retrieved |
|---|---|---|
| #1 | fabaceae OR (legumes or beans or pulses) OR lentil* OR lentils OR *pea* OR vigna* OR bengal gram OR glycine max OR soybean | 233,541 |
| #2 | cholestasis OR cholecystectomy OR cholecystitis OR acalculous cholecystitis OR (choledocholithiasis or cbd stone or common bile duct stones) OR (cholelithiasis or gallstones) OR liver cirrhosis OR biliary tract OR gallstone* OR gallbladder disease OR bile duct stone OR gallbladder inflammation OR gallbladder removal OR acalculous gangrenous cholecystitis OR biliary colic OR biliary calculi OR biliary calculosis OR (cholelithiasis or gallstones) | 32,283 |
| #3 | (non alcoholic fatty liver disease or nafld or fatty liver disease or liver cirrhosis) OR gastroenterology OR non-alcoholic steatohepatitis OR non-alcoholic steatosis OR non alcoholic steatotic hepatitis OR nash cirrhosis OR nash OR liver | 122,133 |
| #4 | #2 OR #3 | 137,811 |
| #5 | 1# AND #4 | 3858 |
| | Limited to dates from 1000/1/1 to 2023/6/30 | |

CINAHL complete (EBSCO Information Services, MA, USA)
Search conducted on July 24th, 2023.

| Search | Query | Records retrieved |
|---|---|---|

| | | |
|---|---|---|
| #1 | fabaceae OR (legumes or beans or pulses) OR lentil* OR lentils OR *pea* OR vigna* OR bengal gram OR glycine max OR soybean | 233,541 |
| #2 | metabolic dysfunction-associated steatotic liver disease OR mafld OR metabolic dysfunction-associated fatty liver disease OR metabolic dysfunction-associated steatohepatitis OR mash | 591 |
| #3 | #1 AND #2 | 21 |
| | Limited to dates from 1000/1/1 to 2023/6/30 | |

Web of Science (Clarivate Analytics, PA, USA)

Search conducted on July 24th, 2023.

| Search | Query | Records retrieved |
|---|---|---|
| #1 | (((((((((((((ALL=(fabaceae)) OR ALL=(lens plant)) OR ALL=(pea*)) OR ALL=(legume*)) OR ALL=(dietary pulse*)) OR ALL=(legume pod)) OR ALL=(lentil*)) OR ALL=(bean*)) OR ALL=(chickpea*)) OR ALL=(vigna)) OR ALL=(phaseolus)) OR ALL=(black gram)) OR ALL=(bengal gram)) OR ALL=(soybean)) OR ALL=(glycine max) | 2,179,533 |
| #2 | ((((((((((((((((ALL=(cholestasis)) OR ALL=(cholelithias*)) OR ALL=(cholecystectomy)) OR ALL=(cholecystitis)) OR ALL=(acalculous cholecystitits)) OR ALL=(choledolithias*)) OR ALL=(liver cirrhosis)) OR ALL=(biliary tract)) OR ALL=(gallstone*)) OR ALL=(gallbladder disease*)) OR ALL=(bile duct gallstone*)) OR ALL=(common bile duct stone*)) OR ALL=(common bile duct gallstone*)) OR ALL=(gallbladder inflammation)) OR ALL=(acalculous gangrene cholecystitis)) OR ALL=(gallbladder removal)) OR ALL=(biliary colic)) OR ALL=(biliary calculi) | 193,391 |
| #3 | (((((((((ALL=(non-alcoholic fatty liver disease)) OR ALL=(nafld)) OR ALL=(non alcoholic fatty liver disease)) OR ALL=(non-alcoholic steatotic hepatitis)) OR ALL=(non alcoholic steatotic hepatitis)) OR ALL=(NASH)) OR ALL=(non-alcoholic stetatohepatitis)) OR ALL=(non alcoholic steatohepatitis)) OR ALL=(non-alcoholic fatty liver)) OR ALL=(non alcoholic fatty liver) | 104,775 |
| #4 | #2 OR #3 | 290,922 |
| #5 | #1 AND #4 | 7365 |
| | Limited to dates from 1000/1/1 to 2023/6/30 | |

Web of Science (Clarivate Analytics, PA, USA)

Search conducted on July 24th, 2023.

| Search | Query | Records retrieved |
|---|---|---|
| #1 | (((((((((((((ALL=(fabaceae)) OR ALL=(lens plant)) OR ALL=(pea*)) OR ALL=(legume*)) OR ALL=(dietary pulse*)) OR ALL=(legume pod)) OR ALL=(lentil*)) OR ALL=(bean*)) OR ALL=(chickpea*)) OR ALL=(vigna)) OR ALL=(phaseolus)) OR ALL=(black gram)) OR ALL=(bengal gram)) OR ALL=(soybean)) OR ALL=(glycine max) | 2,179,533 |
| #2 | (((ALL=(metabolic dysfunction-associated fatty liver disease)) OR ALL=(metabolic dysfunction-associated steatohepatitis)) OR ALL=(MAFLD)) OR ALL=(MASH) | 9852 |
| #3 | #1 AND #2 | 804 |
| | Limited to dates from 1000/1/1 to 2023/6/30 | |

### Appendix II: Search strategy for grey and unpublished literature

ProQuest (Clarivate Analytics, PA, USA)

Search conducted on July 24th, 2023.

| Search | Query | Records retrieved |
|---|---|---|
| S1 | (Fabaceae OR fabaceae OR (lens plant) OR lentil OR pea OR legume OR pulses OR (legume pod) OR bean OR vigna) | 2,994,928 |
| S2 | ((phaseolus vulgaris) OR (phaseolus mungo) OR (black gram) OR (red gram) OR chickpea OR (bengal gram) OR phaseolus OR soybean OR soy OR (glycine max)) | 1,848,248 |
| S3 | (cholestasis OR cholecystectomy OR cholecystitis OR (acalculous cholecystitis) OR choledocholithiasis OR cholelithiasis OR (liver cirrhosis) OR (biliary tract) OR gallstone OR (gallbladder disease)) | 212,048 |
| S4 | ((bile duct gallstone) OR (common bile duct gallstone) OR (gallbladder inflammation) OR (gallbladder inflammatory) OR (acalculous cholecystitis) OR (acalculous gangrenous cholecystitis) OR cholelithias OR choledocholithiasis OR choledocholithias OR cholecystectomy) | 34,046 |
| S5 | (cholecystectomies OR (gallbladder removal) OR (removal of gallbladder) OR (biliary colic) OR (biliary calculi) OR (biliary calculosis)) | 26,540 |
| S6 | ((Non-alcoholic Fatty Liver Disease) OR gastroenterology OR nafld OR nash OR (non alcoholic fatty liver) OR (non alcoholic fatty liver disease) OR (non alcoholic steatotic hepatitis) OR (non alcoholic steotohepatitis)) | 803,805 |
| S7 | [S1] OR [S2] | 4,524,753 |
| S8 | [S3] OR [S4] OR [S5] OR [S6] | 938,769 |
| S9 | [S7] AND [S8] | 116,647 |
| S10 | [S9] AND pd(20220601-20230630) | 15,319 |
| S11 | mice OR rats OR rat OR chicken OR broiler OR dog OR hamster OR cat OR rabbit OR broilers OR hamsters OR mouse OR animal OR lama OR alpaca | 11,168,079 |
| S12 | [S10] NOT [S11] | 3667 |
| | Limited to dates from 2022/6/1 to 2023/6/30 | |

Google scholar (Google Inc., CA, USA)
Search conducted on July 24$^{th}$, 2023.

| Search | Query | Records retrieved |
|---|---|---|
| #1 | (fabaceae OR legumes OR pulses OR lentils OR bean) AND (non-alcoholic fatty liver OR non-alcoholic steatotic hepatitis OR gallstone OR gallbladder disease OR cholecystectomy OR removal of gallbladder OR cholelithiasis) | 141 |
| | Limited to dates from 2022/6/1 to 2023/6/30 | |

#### Appendix III: Studies ineligible following full text review

| | Studies from databases and registries | |
|---|---|---|
| # | Article | Reason for exclusion |
| 1 | Adriano, L. S., De Carvalho Sampaio, H. A., Arruda, S. P. M., De Melo Portela, C. L., De Melo, M. L. P., Carioca, A. A. F., & Soares, N. T. (2016). Healthy dietary pattern is inversely associated with non-alcoholic fatty liver disease in elderly [Article]. *British Journal of Nutrition*, *115*(12), 2189-2195. https://doi.org/10.1017/S0007114516001410 | Wrong exposure; not legume as a separate exposure |
| 2 | Alferink, L. J. M., Erler, N. S., de Knegt, R. J., Janssen, H. L. A., Metselaar, H. J., Darwish Murad, S., & Kiefte-de Jong, J. C. (2020). Adherence to a plant-based, high-fibre dietary pattern is related to regression of non-alcoholic fatty liver disease in an elderly population [Article]. *European Journal of Epidemiology*, *35*(11), 1069-1085. https://doi.org/10.1007/s10654-020-00627-2 | Wrong exposure; not legume as a separate exposure |
| 3 | Carr, T. (2001). Soy & health 2000: Clinical evidence, dietetic applications [Conference Paper]. *Nutrition Bulletin*, *26*(2), 179-182. | Wrong outcome; does not investigate hepatobiliary diseases. |
| 4 | Duane, W. C. (1997). Effects of legume consumption on serum cholesterol, biliary lipids, and sterol metabolism in humans. *Journal of lipid research*, *38*(6), 1120-1128. | Wrong outcomes; only evaluates cholesterol mechanisms. |
| 5 | Ferolla, S. M., Ferrari, T. C., Lima, M. L., Reis, T. O., Tavares, W. C., Jr., Couto, O. F., Vidigal, P. V., Fausto, M. A., & Couto, C. A. (2013). Dietary patterns in Brazilian patients with nonalcoholic fatty liver disease: a cross-sectional study. *Clinics (Sao Paulo)*, *68*(1), 11-17. https://doi.org/10.6061/clinics/2013(01)oa03 | Wrong exposure; not legume as a separate exposure |
| 6 | Gamonski, W. (2014). The Mighty Mung Bean. *Life Extension*, 89-84. | Wrong outcome; does not investigate hepatobiliary diseases. |
| 7 | Iriyama, H., Kato, M., Nogami, M., & Tokuda, Y. (2014). Edamame (green soy beans) biliary stones. *BMJ Case Rep*, *2014*. https://doi.org/10.1136/bcr-2014-207677 | Wrong exposure; no mention of legumes, is a case report of a woman with a bean-shaped gallstone. |

| | | |
|---|---|---|
| 8 | Juárez-Chairez, M. F., Meza-Márquez, O. G., Márquez-Flores, Y. K., & Jiménez-Martínez, C. (2022). Potential anti-inflammatory effects of legumes: a review [Article]. *British Journal of Nutrition*, *128*(11), 2158-2169. https://doi.org/10.1017/S0007114522000137 | Wrong exposure; not legume as a separate exposure |
| 9 | Kouvari, M., Tsiampalis, T., Chrysohoou, C., Georgousopoulou, E., Skoumas, J., Mantzoros, C. S., Pitsavos, C., & Panagiotakos, D. B. (2021). The quality of plant-based dietary patterns affects the ten-year cardiovascular disease risk of participants with non-alcoholic fatty liver disease: Highlights from a population-based cohort study [Conference Abstract]. *European Heart Journal*, *42*(SUPPL 1), 2432. https://doi.org/10.1093/eurheartj/ehab724.2432 | Wrong exposure; not legume as a separate exposure |
| 10 | Kron, J. (2008). Cholelithiasis. *JOURNAL OF COMPLEMENTARY MEDICINE*, *7*(3), 22-26. | Wrong population; refer to animal studies for the associations within the scope of the review. |
| 11 | Li, Y., Deng, X., Guo, X., Zhang, F., Wu, H., Qin, X., & Ma, X. (2023). Preclinical and clinical evidence for the treatment of non-alcoholic fatty liver disease with soybean: A systematic review and meta-analysis [Review]. *FRONTIERS IN PHARMACOLOGY*, *14*. https://doi.org/10.3389/fphar.2023.1088614 | Wrong intervention; investigates treatment with soy among individuals with prevalent non-alcoholic fatty liver disease |
| 12 | Liu, X., Peng, Y., Chen, S., & Sun, Q. (2018). An observational study on the association between major dietary patterns and non-alcoholic fatty liver disease in Chinese adolescents [Article]. *Medicine (United States)*, *97*(17). https://doi.org/10.1097/MD.0000000000010576 | Wrong exposure; not legume as a separate exposure |
| 13 | Matsumoto, Y., Fujii, H., Harima, M., Okamura, H., Yukawa-Muto, Y., Odagiri, N., Motoyama, H., Kotani, K., Kozuka, R., Kawamura, E., Hagihara, A., Uchida-Kobayashi, S., Enomoto, M., Yasui, Y., Habu, D., & Kawada, N. (2023). Severity of Liver Fibrosis Is Associated with the Japanese Diet Pattern and Skeletal Muscle Mass in Patients with Nonalcoholic Fatty Liver Disease. *Nutrients*, *15*(5), Article 1175. https://doi.org/10.3390/nu15051175 | Wrong exposure; not legume as a separate exposure |
| 14 | Miryan, M., Darbandi, M., Moradi, M., Najafi, F., Soleimani, D., & Pasdar, Y. (2023). Relationship between the Mediterranean diet and risk of hepatic fibrosis in patients with non-alcoholic fatty liver disease: A cross-sectional analysis of the RaNCD cohort. *Front Nutr*, *10*, 1062008. https://doi.org/10.3389/fnut.2023.1062008 | Wrong exposure; not legume as a separate exposure |
| 15 | Montemayor, S., Bouzas, C., Mascaro, C. M., Casares, M., Llompart, I., Abete, I., Angullo-Martinez, E., Zulet, M. A., Martinez, J. A., & Tur, J. A. (2022). Effect of Dietary and Lifestyle Interventions on the Amelioration of NAFLD in | Wrong exposure; not legume as a separate exposure |

| | | |
|---|---|---|
| | Patients with Metabolic Syndrome: The FLIPAN Study. *Nutrients*, *14*(11), Article 2223. https://doi.org/10.3390/nu14112223 | |
| 16 | Nakashita, C., Xi, L., Inoue, Y., Kabura, R., Masuda, S., Yamano, Y., & Katoh, T. (2021). Impact of dietary compositions and patterns on the prevalence of nonalcoholic fatty liver disease in Japanese men: a cross-sectional study. *BMC GASTROENTEROLOGY*, *21*(1), 1-8. https://doi.org/10.1186/s12876-021-01919-x | Wrong exposure; not legume as a separate exposure |
| 17 | Naseri, K., Saadati, S., Asadzadeh-Aghdaei, H., Hekmatdoost, A., Sadeghi, A., Sobhani, S. R., Abhari, K., Bahrami, A., Rahimi-Sakak, F., Jamshidfar, N., & Zali, M. (2022). Healthy Dietary Pattern Reduces Risk of Gallstones: Results of a Case-Control Study in Iran. *International Journal of Preventive Medicine*, *13*(1), Article 66. https://doi.org/10.4103/ijpvm.IJPVM_455_19 | Wrong exposure; not legume as a separate exposure |
| 18 | Nervi, F., Covarrubias, C., Bravo, P., Velasco, N., Ulloa, N., Cruz, F., Fava, M., Severín, C., Del Pozo, R., Antezana, C., & et al. (1989). Influence of legume intake on biliary lipids and cholesterol saturation in young Chilean men. Identification of a dietary risk factor for cholesterol gallstone formation in a highly prevalent area. *Gastroenterology*, *96*(3), 825-830. | Wrong outcome; investigates cholesterol composition in bile and blood, not gallbladder disease. |
| 19 | Salehi-sahlabadi, A., Teymoori, F., Jabbari, M., Momeni, A., Mokari-yamchi, A., Sohouli, M., & Hekmatdoost, A. (2021). Dietary polyphenols and the odds of non-alcoholic fatty liver disease: A case-control study [Article]. *Clinical Nutrition ESPEN*, *41*, 429-435. https://doi.org/10.1016/j.clnesp.2020.09.028 | Wrong exposure; not legume as a separate exposure |
| 20 | Sayegh, N. F., Heraoui, G. N., Sayegh, L. N., & Sayegh, R. (2020). Relation of dietary patterns and nutritional profile to hepatic fibrosis in a sample of Lebanese non-alcoholic fatty liver disease patients [Conference Abstract]. *Obesity Reviews*, *21*(SUPPL 1). https://doi.org/10.1111/obr.13118 | Wrong exposure; not legume as a separate exposure |
| 21 | Shanmugam, H. (2022). Mediterranean diet: food for changing the NAFLD world [Conference Abstract]. *European Journal of Clinical Investigation*, *52*, 27. https://doi.org/10.1111/eci.13796 | Wrong exposure; not legume as a separate exposure |
| 22 | Shanmugam, H., Molina, E., Di Palo, D. M., Di Ciaula, A., & Portincasa, P. (2020). Adherence to Mediterranean diet in southern Italy. older do better than younger subjects [Conference Abstract]. *European Journal of Clinical Investigation*, *50*(SUPPL 1), 16. https://doi.org/10.1111/eci.13369 | Wrong exposure; not legume as a separate exposure |

| | | |
|---|---|---|
| 23 | Spathis, A., Heaton, K. W., Emmett, P. M., Norboo, T., & Hunt, L. (1997). Gallstones in a community free of obesity but prone to slow intestinal transit. *Eur J Gastroenterol Hepatol*, *9*(2), 201-206. https://doi.org/10.1097/00042737-199702000-00018 | Wrong exposure; not legume as a separate exposure |
| 24 | Sun, P., Huang, L. P., Shuai, P., Wan, Z. W., Liu, Y. Y., Xue, J. Q., & Liu, Y. P. (2022). Effect of a High Protein, Low Glycemic Index Dietary Intervention on Metabolic Dysfunction-Associated Fatty Liver Disease: A Randomized Controlled Trial. *FRONTIERS IN NUTRITION*, *9*, Article 863834. https://doi.org/10.3389/fnut.2022.863834 | Wrong exposure; not legume as a separate exposure |
| 25 | van Eekelen, E., Geelen, A., Alssema, M., Lamb, H. J., de Roos, A., Rosendaal, F. R., & de Mutsert, R. (2020). Adherence to dietary guidelines in relation to visceral fat and liver fat in middle-aged men and women: the NEO study [Article]. *International Journal of Obesity*, *44*(2), 297-306. https://doi.org/10.1038/s41366-019-0441-x | Wrong exposure; not legume as a separate exposure |
| 26 | Vieira, N. M., Peghinelli, V. V., Monte, M. G., Costa, N. A., Pereira, A. G., Seki, M. M., Azevedo, P. S., Polegato, B. F., de Paiva, S. A. R., Zornoff, L. A. M., & Minicucci, M. F. (2023). Beans consumption can contribute to the prevention of cardiovascular disease [Review]. *Clinical Nutrition ESPEN*, *54*, 73-80. https://doi.org/10.1016/j.clnesp.2023.01.007 | Wrong outcome; not metabolic dysfunction-associated fatty liver disease or gallbladder disease. |
| 27 | Vitale, M., Della Pepa, G., Costabile, G., Bozzetto, L., Cipriano, P., Signorini, S., Leoni, V., Riccardi, G., Vaccaro, O., & Masulli, M. (2022). Association between Diet Quality and Index of Non-Alcoholic Steatohepatitis in a Large Population of People with Type 2 Diabetes: Data from the TOSCA.IT Study. *Nutrients*, *14*(24), 5339. https://doi.org/10.3390/nu14245339 | Wrong exposure; not legume as a separate exposure |
| 28 | Xiong, P., & Zhu, Y. F. (2021). Soy diet for nonalcoholic fatty liver disease: A meta-analysis of randomized controlled trials. *Medicine (Baltimore)*, *100*(22), e25817. https://doi.org/10.1097/md.0000000000025817 | Wrong outcome; investigates the association between soy and co-morbidities among individuals with prevalent metabolic dysfunction-associated fatty liver disease. |
| 29 | Yao, Y., Hao, L., Shi, Z., Wang, L., Cheng, X., Wang, S., & Ren, G. (2014). Mung bean decreases plasma cholesterol by up-regulation of CYP7A1. *Plant Foods Hum Nutr*, *69*(2), 134-136. https://doi.org/10.1007/s11130-014-0405-1 | Wrong population; study on hamsters |
| 30 | Yip, C. S. C., Yip, Y. C., & Chan, W. (2023). The associations of soya intakes with non-communicable diseases: a scoping review of meta-analyses [Article]. *British Journal of Nutrition*, *129*(1), 135-146. https://doi.org/10.1017/S0007114522000691 | Wrong outcome; does not investigate metabolic dysfunction- |

| | | |
|---|---|---|
| | | associated fatty liver disease or gallbladder disease. |
| 31 | Zarei, A., Stasi, C., Mahmoodi, M., Masoumi, S. J., Zare, M., & Jalali, M. (2020). Effect of soy consumption on liver enzymes, lipid profile, anthropometry indices, and oxidative stress in patients with non-alcoholic fatty liver disease: A systematic review and meta-analysis of clinical trials [Article]. *IRANIAN JOURNAL OF BASIC MEDICAL SCIENCES*, *23*(10), 1245-1250. https://doi.org/10.22038/ijbms.2020.46854.10797 | Wrong outcome; investigates the association between soy and co-morbidities among individuals with prevalent metabolic dysfunction-associated fatty liver disease. |
| 32 | Zhang, W., Wang, X. Y., Huang, J. L., Wang, S. Y., Yao, Q., & Li, H. W. (2023). Healthy Eating Index-2015 in relation to risk of metabolic dysfunction-associated fatty liver disease among US population: National Health and Nutrition Examination Survey 2017-2018. *FRONTIERS IN NUTRITION*, *9*, Article 1043901. https://doi.org/10.3389/fnut.2022.1043901 | Wrong exposure; not legume as a separate exposure |
| 33 | CME Exam 3: Improved Diet Quality Associates With Reduction in Liver Fat, Particularly in Individuals With High Genetic Risk Scores for Nonalcoholic Fatty Liver Disease. (2018). [Note]. *Gastroenterology*, *155*(1), e25-e26. https://doi.org/10.1053/j.gastro.2018.06.026 | Wrong intervention; gastroenterology exam |
| | Studies identified via other methods | |
| # | Article | Reason for exclusion |
| 1 | Ling, Z., Zhang, C., He, J., Ouyang, F., Qiu, D., Li, L., Li, Y., Li, X., Duan, Y., Luo, D., Xiao, S., & Shen, M. (2023). Association of Healthy Lifestyles with Non-Alcoholic Fatty Liver Disease: A Prospective Cohort Study in Chinese Government Employees. *Nutrients*, *15*(3). https://doi.org/10.3390/nu15030604 | Wrong exposure; not legume as a separate exposure |
| 2 | Jahromi, M. K., Daftari, G., Farhadnejad, H., Tehrani, A. N., Teymoori, F., Salehi-Sahlabadi, A., & Mirmiran, P. (2023). The association of healthy lifestyle score and risk of non-alcoholic fatty liver disease. *BMC public health*, *23*(1), 973. https://doi.org/10.1186/s12889-023-15816-3 | Wrong exposure; not legume as a separate exposure |
| 3 | Torres-Peña, J. D., Arenas-de Larriva, A. P., Alcala-Diaz, J. F., Lopez-Miranda, J., & Delgado-Lista, J. (2023). Different Dietary Approaches, Non-Alcoholic Fatty Liver Disease and Cardiovascular Disease: A Literature Review. *Nutrients*, *15*(6), 1483. https://doi.org/10.3390/nu15061483 | Wrong exposure; not legume as a separate exposure |

| | | |
|---|---|---|
| 4 | Korpijaakko, C. A., Eriksson, J. G., Kautiainen, H., Klemetti, M. M., & Laine, M. K. (2023). Fatty liver index in young adult offspring of women with type 1 diabetes. *Diabetology & Metabolic Syndrome*, *15*(1), 196. https://doi.org/10.1186/s13098-023-01164-0 | Wrong exposure; did not evaluate legumes as exposure. |
| 5 | Spiezia, C., Di Rosa, C., Fintini, D., Ferrara, P., De Gara, L., & Khazrai, Y. M. (2023). Nutritional Approaches in Children with Overweight or Obesity and Hepatic Steatosis. *Nutrients*, *15*(11), 2435. https://www.mdpi.com/2072-6643/15/11/2435 | Wrong exposure; not legume as a separate exposure |
| 6 | Jakobek, L., & Blesso, C. Beneficial effects of phenolic compounds: native phenolic compounds vs metabolites and catabolites. *Critical Reviews in Food Science and Nutrition*, 1-19. https://doi.org/10.1080/10408398.2023.2208218 | Wrong exposure; phenolic compounds found in legumes. |
| 7 | D'Aoust, L., & Poitras, P. (2022). Diets and Digestive Diseases. In P. Poitras, M. Bilodeau, M. Bouin, & J.-E. Ghia (Eds.), *The Digestive System: From Basic Sciences to Clinical Practice* (pp. 417-424). Springer International Publishing. https://doi.org/10.1007/978-3-030-98381-9_29 | Wrong comparator; does not evaluate the association between legumes and hepatobiliary health. |
| 8 | Antunes, C., Arbo, M. D., & Konrath, E. L. (2022). Hepatoprotective Native Plants Documented in Brazilian Traditional Medicine Literature: Current Knowledge and Prospects. *Chemistry & biodiversity*, *19*(6), e202100933-n/a. https://doi.org/10.1002/cbdv.202100933 | Wrong exposure; investigates legume extracts only. |
| 9 | Foghis, M., Bungau, S. G., Bungau, A. F., Vesa, C. M., Purza, A. L., Tarce, A. G., Tit, D. M., Pallag, A., Behl, T., ul Hassan, S. S., & Radu, A.-F. (2023). Plants-based medicine implication in the evolution of chronic liver diseases. *BIOMEDICINE & PHARMACOTHERAPY*, *158*, 114207. https://doi.org/https://doi.org/10.1016/j.biopha.2022.114207 | Wrong exposure; investigates legume extracts only. |
| 10 | Ray Sarkar, B., & Das, J. (2023). A REVIEW OF ETHNOMEDICINAL PLANTS AND THEIR TRADITIONAL USES FOR THE TREATMENT OF HEPATOBILIARY DISEASE BY DIFFERENT TRIBES OF NORTH-EAST INDIA. | Wrong exposure; investigates a tree in the fabaceae species and not legumes. |
| 11 | Maheshwari, S., Kumar, S., Nakshiwala, B. V., Srivastav, A., Chavan, V., Raut, A., & Maheshwari, A. (2022). Fatty Liver Disease: Pathophysiology and Imaging Features. *Indographics*, *01*(01), 057-077. https://doi.org/10.1055/s-0042-1742574 | Wrong exposure; does not evaluate legumes or other dietary factors in relation to hepatobiliary diseases. |
| 12 | Soltanieh, S., Salavatizadeh, M., Poustchi, H., Yari, Z., Mansour, A., Khamseh, M. E., Malek, M., Alaei-Shahmiri, F., & Hekmatdoost, A. (2023). The association of dietary inflammatory index (DII) and central obesity with non-alcoholic fatty | Wrong exposure; not legume as a separate exposure |

| | | |
|---|---|---|
| | liver disease (NAFLD) in people with diabetes (T2DM). *Heliyon*, *9*(3), e13983. https://doi.org/10.1016/j.heliyon.2023.e13983 | |
| 13 | Fu, J., & Shin, S. (2023). Dietary patterns and risk of non-alcoholic fatty liver disease in Korean adults: a prospective cohort study. *BMJ Open*, *13*(1), e065198. https://doi.org/10.1136/bmjopen-2022-065198 | Wrong exposure; not legume as a separate exposure |
| 14 | Munteanu, C., & Schwartz, B. (2023). The Effect of Bioactive Aliment Compounds and Micronutrients on Non-Alcoholic Fatty Liver Disease. *ANTIOXIDANTS*, *12*(4). https://doi.org/10.3390/antiox12040903 | Wrong exposure; only evaluates cocoa beans and peanuts |

#### Appendix IV: Data extraction instrument

| Author, study aim and study type | Population and context | Concept | Statistical methods<br><br>For non-evidence articles this might be irrelevant. | Confounding or other bias<br><br>For non-evidence article: which potential confounders are presented/discussed? | Key findings |
|---|---|---|---|---|---|
| First author et al. (reference)<br><br>Evidence level articles: Aim<br><br>Non-evidence and narrative reviews: Theme Study design or article type | Number of participants, drop-out rates or % participation.<br><br>Specific information on sex, age, etc.<br><br>Which population/setting are participants recruited from. | Description of exposure, outcome, duration of study. Short description of intervention | Information about statistical modelling and/or analyses conducted | Description of how confounding or other potential biases was dealt with, e.g., loss to follow-up (comparison of participants and non-participants), validated methods, etc.<br><br>For textbooks and expert text opinion papers, include the source of information here | Description of key findings in relation to the focus of this review. Presented as estimates and SD/SE/CI or text if estimates not applicable.<br><br>Tables displayed in this column closely resemble or are substantially derived from tables found in the original primary article from which the data extraction was sourced. |

### Appendix V: Methodological quality assessment of included studies

#### *Cohort studies*

*Quality assessment of included cohort studies with Scottish Intercollegiate Guidelines Network checklist (SIGN).* [1]

| Author | 1.1. Appropriate and clearly focused question | 1.2. The two study groups are comparable in all respects other than the factor under investigation | 1.3. Indication of participation percentage | 1.4. Likelihood that eligible subjects have the outcome at enrolment | 1.5. Percentage dropout before study completion | 1.6. Comparison between full participants and drop-outs | 1.7. Outcomes are clearly defined | 1.8. Blinded outcome assessment | 1.9. If non-blinded outcome assessment; is knowledge on exposure accounted for | 1.10. The method for assessment of exposure is reliable | 1.11. Other sources used to validate and assess reliability of methods for outcome assessment. | 1.12. Exposure level assessed more than once | 1.13. Potential confounders identified and accounted for | 1.14. Provision of confidence intervals | 2.1. Minimized risk of bias or confounding | 2.2. Clear evidence of association between exposure and outcome | 2.3. Results directly applicable to target patient group | 2.4. Notes. |
|---|---|---|---|---|---|---|---|---|---|---|---|---|---|---|---|---|---|---|
| Barré et al. (2017) [2] | √ | √ | √ | √ | 3.19 | √ | √ | √ | √ | √ | √ | ÷ | √ | √ | ++ | √ | √ | Author conclusions: There was an inverse dose-response relationship between legumes and cholecystectomy.<br><br>The article examines women insured through teaching plans, comparing high and low legume consumption. Adjustments were made for various factors like BMI, surgery history, and education. Around 100,000 out of 500,000 invited women participated, [3] with 13.4% excluded, but the excluded groups' legume consumption remains unknown. A flow chart or missing data details in tables would enhance clarity. Individuals with prior cholecystectomy were excluded from the study at baseline. About 3.19% were lost since baseline. The article refers to another article where completers and non-completers are compared. Self-reported outcomes were validated within the cohort, while exposure was also self-reported and known during outcome assessments. Any misclassification would likely bias towards the null. The interaction between dietary pattern and covariables was tested to ensure that exposure assessment was accounted for as the participants were non-blinded. Exposure data was collected only at baseline, with adjustments and imputations made for missing data. Confidence intervals were provided. The risk of bias seems minimal, yet potential bias could bias associations toward null. |
| Tsai et al. (2006) [4] | √ | √ | √ | ? | 20 | ÷ | √ | ÷ | ÷ | √ | √ | √ | √ | √ | + | ÷ | √ | Author conclusions: No significant protective association between legume consumption and cholecystectomy was found.<br><br>Adjustments are made in analyses to account for group differences in various quintiles of diet intakes. Participation rate stands at approximately 63%, with exclusions due to inadequate data for the study's aim. Over 90% completed subsequent biennial questionnaires, and 80% filled out repeated dietary questionnaires during follow-up. Self-reported gallbladder removal is used as a proxy for gallstones, validated within the cohort. The study assumes awareness of surgical organ removal, querying cholecystectomy reports from 1980 onwards. Participant flow isn't illustrated, and non-participants aren't detailed. The reliance on self-reports for outcomes and exposures results in potential biases. Diet information is gathered multiple times, adjusting exposure to reflect dietary changes, and validated accordingly. Findings show a non-significant association between legumes and cholecystectomy. |
| Zhang et al. (2023) [5] | √ | √ | ? | √ | TCLSIH: 11.7<br><br>GNHS: 19.2 | ÷ | √ | ÷ | ÷ | √ | √ | √ | √ | √ | ++ | ? | √ | Author conclusions: Replacing animal protein foods with plant protein foods is related to a significant reduction in NAFLD risk.<br><br>Participants are drawn from large national cohorts in China, aiming to represent the general Chinese population. The studies include all individuals hospitalized within the baseline period, potentially accessing information from insurance registries covering nearly all hospital visits in China. This should have been mentioned in the article if relevant.<br><br>Those with liver diseases at baseline were excluded. There's no comparison between partial or full comparisons. Blinding of outcome assessments isn't mentioned. Diet assessment involved validated FFQs and analytical models, conducted yearly in TCLSIH and every third year in GNHS. Misclassification of NAFLD presence is likely low due to professional assessment. Any bias would likely lean towards null effects. Efforts to address confounding and bias have been made. |

| | | | | | | | | | | | | | | | | | |
|---|---|---|---|---|---|---|---|---|---|---|---|---|---|---|---|---|---|
| | | | | | | | | | | | | | | | | | Regarding legume consumption, a reduced HR for NAFLD was observed when substituting with poultry in GNHS and borderline significance when substituting with processed meats in the TCLSIH cohort. The scoping review's PICO covers all global populations, making the results applicable to the review's target population |

*Yes (√), No (÷), Unknown (?), High quality (++), Acceptable (+), Unacceptable (0). TCLSIH, Tianjin Chronic Low-grade Systemic Inflammation and Health. GNHS, Guangzhou Nutrition and Health Study. NAFLD, non-alcoholic fatty liver disease. FFQ, food frequency questionnaire. HR, hazard ratio,*

#### Case-control studies

*Quality assessment of included case-control studies with Scottish Intercollegiate Guidelines Network checklist (SIGN).* [1]

| | 1.1. Appropriate and clearly focused question | 1.2. The two study groups are comparable | 1.3. The same exclusion criteria are used for both cases and controls. | 1.4. Percentage of each group (cases/controls) participated in the study | 1.5. Comparison between full participants and drop-outs. | 1.6. Cases are clearly defined and differentiated from controls. | 1.7. It is clearly established that controls are non-cases | 1.8. Limited knowledge of primary exposure influencing case ascertainment | 1.9. Exposure and outcome status is measured in a standard, valid and reliable way | 1.10. Potential confounders identified and accounted for | 1.11. Provision of confidence intervals | 2.1. Minimized risk of bias or confounding | 2.2. Clear evidence of association between exposure and outcome | 2.3. Results directly applicable to target patient group | 2.4. Notes. |
|---|---|---|---|---|---|---|---|---|---|---|---|---|---|---|---|
| Bahrami et al. (2019) [6] | √ | √ | √ | 94/ 98 | ÷ | √ | √ | ? | √ | √ | √ | + + | √ | √ | Author conclusions: Higher consumption of beans, lentils, and peas was associated with lower risk of NAFLD.<br><br>Cases and controls are all referred to a gastroenterology clinic. Both groups have the same exclusion criteria but only based on energy intake, potentially limiting the transferability of the study as it provides minimal information on the criteria for inclusion or exclusion. There's high participation, but this information is not included in their own study; instead, it refers to another study/protocol. There's no information provided regarding blinding. Risk of bias minimal. |
| Giraldi et al. (2020) [7] | √ | √ | ? | ÷ | ÷ | √ | ? | √ | ? | √ | √ | + | ? | √ | Author conclusions: Highest compared to lowest legume consumption was protective of NAFLD.<br><br>The study focuses on diet patterns, with relevant sub-analyses for the review. Cases with NAFLD and controls without NAFLD were selected from the same hospitals, but the exclusion criteria for controls were not clearly specified. Details on participation rates and non-participants are missing, and the method for excluding NAFLD among controls isn't specified. NAFLD was diagnosed following validated standard measures from the American Gastroenterology Association. Diagnosis preceded the diet assessment, which used a validated FFQ specifically for the Mediterranean Diet. There was no description of the validation in the text or in the referenced article for the validation statement and exposure assessment quality is thus unknown. Confounders were adjusted for, and CIs were provided. There's a potential risk of selection bias due to unclear selection criteria, potentially affecting participants' knowledge of their outcomes while reporting their diets. Thorough data collection might alter association magnitudes. |
| Han et al. (2014) [8] | √ | √ | √ | ÷ | ÷ | √ | ? | √ | ? | √ | √ | 0 | ÷ | √ | Author conclusions: There was no association between bean consumption and NAFLD.<br><br>Cases and controls are recruited from the same population, with relatively similar exclusion and inclusion criteria for both cases and controls. It's not entirely clear that controls are not cases, as there's no mention of exclusion due to NAFLD, except that controls underwent an ultrasound scan within three months. NAFLD is measured before exposure information is collected. There aren't specific details about the NAFLD diagnosis beyond the use of ultrasound, lacking information on diagnostic criteria, etc. Dietary assessment is done through 24-hour recall and food diaries, both commonly used methods, but there's no internal validation or reference to other literature, making it uncertain how well-suited these tools are in this context. It's uncertain to what extent bias exists due to selection and information since these are not sufficiently described. It's unclear if controls might have NAFLD, which may result in the comparison groups becoming too similar, potentially obscuring any differences. There's a lack of information to assess selection bias. |
| Hao et al. (2021) [9] | √ | ? | √ | ÷ | ÷ | √ | √ | ? | √ | ÷ | √ | 0 | ÷ | √ | Author conclusions: The influence of bean products on OR for NAFLD was marginally different between carriers of the C and the T allele.<br><br>They haven't provided specific dropout rates for cases and controls separately, but only 1% of participants were excluded. The total number of individuals invited to take part remains undisclosed. Cases in the study have been diagnosed with NAFLD, and controls are excluded based on this criterion to ensure they aren't misclassified into the wrong outcome group, but there is no mention of matching criteria between cases and controls, and the baseline tables display some differences in characteristics between cases and controls. Moreover, all participants underwent ultrasound scans. There's uncertainty about |

| | | | | | | | | | | | | | | | |
|---|---|---|---|---|---|---|---|---|---|---|---|---|---|---|---|
| | | | | | | | | | | | | | | | whether exposure assessment preceded the outcome evaluation, although the outcome assessment was conducted by an expert. Both exposure and outcome assessments were conducted in a standardized and validated manner for this population. Additionally, the dietary assessment used has been validated within the Chinese population. While the study adjusts for age, it's not clear if other factors are also considered. Despite identifying numerous confounders, they haven't accounted for them in the analysis, and the selection process lacks clarity, potentially impacting the accuracy of the association estimate. This could introduce considerable bias, leading to different outcomes if the analyses were redone. An association between legumes and NAFLD exists, but the size and direction of this association might be influenced by unaddressed confounding factors. |
| Jayanthi et al. (2005) [10] | √ | √ | √ | ÷ | ÷ | √ | √ | ÷ | ? | ÷ | √ | 0 | ÷ | √ | Author conclusion: Tamarind consumed more than three times weekly compared to less than three times weekly was associated with increased risk of gallstones.<br><br>There's a lack of information about invitation numbers, dropouts, or exclusions. The cases encompass individuals diagnosed with both symptomatic and asymptomatic gallstones, while controls are included if they have normal ultrasound scans. The sequence of exposure and outcome assessments remains unspecified. The diet assessment only considered frequency, neglecting to measure consumption amounts, potentially affecting the outcome risk. The diagnosis of gallstones relied on ultrasound, the standard method. Although multivariate logistic regression was referenced, there's no clarification on the confounding factors included in the adjustments. The inadequate description of all methods raises significant doubts about the reliability of the estimated associations. If the analyses were conducted again with more detail, it's likely that the resulting associations would undergo substantial changes. |
| Parveen et al. (2019) [11] | √ | ? | ? | ÷ | ÷ | ? | √ | ? | ? | ÷ | √ | 0 | ÷ | √ | Author conclusions: Mung and masoor were positively associated with gallstone.<br><br>Insufficient information exists about controls to assess if cases and controls originate from the same source population. Inclusion and exclusion criteria lack specificity. There are concerns about recall bias regarding various factors and potential residual confounding from multiple variables like age, gender, medication use, obesity, weight loss, fasting, and physical activity, which are poorly explained. The questionnaire used to assess risk factors was designed specifically for this study but lacks validation. The reported positive significant associations between mung beans and masoor pulses are not reflected in the 95% CI, which include 1 for both exposures. The comparison group is not mentioned but appear to be mixed pulses consumed once weekly. There's discrepancy between the stated investigation of daily consumption in the results text and the presentation of consumption once per week as the exposure of the dietary component in the tables. |
| Pastides et al. (1990) [12] | √ | √ | √ | 97/ 97 | ÷ | √ | √ | ÷ | √ | ÷ | ÷ | 0 | ? | √ | Author conclusions: No association between pulses/beans and gallstones<br><br>The lack of information about blinding or independent assessment of exposure and outcome raises uncertainty regarding the knowledge of exposure status at the outcome definition stage. They justify their exposure assessment methods as other studies have used similar methods. |
| Rasheed et al. (2020) [13] | √ | ? | √ | ÷ | ÷ | √ | ? | ÷ | ÷ | ÷ | ÷ | 0 | ? | ÷ | Author conclusion: Legume consumption is higher among cases than controls.<br><br>Controls lack detailed descriptions, raising the possibility of differences in participant characteristics across hospitals. The sampling technique, specifically regarding the index date or similar, isn't clearly explained despite the potential use of incidence density sampling based on gallstone diagnosis criteria. Cases were interviewed using predetermined outcome-specific questions, possibly indicating assessors' awareness of outcome status during exposure assessment. The authors claim validated data collection methods but refer to an article with no diet collection and no mention of validation. There are no statistical adjustments and limited description of matching between cases and controls. CIs are missing from the reported results. While potentially transferable, the study's poor quality renders the results unusable, posing challenges in scoring the applicability of results to the scoping review population. |
| Thijs et al. (1990) [14] | √ | √ | √ | 86/ 72 | ? | √ | √ | ÷ | ÷ | ÷ | ÷ | 0 | √ | √ | Authors conclusion: Regular intake of legumes, 8 or more times monthly, versus eating legumes less than once monthly was associated with a 50% lower OR for gallstones.<br><br>In the acute cases, 14% were non-responders, while the non-responder rate for elective cases and controls is not detailed. Non-participants are mentioned in Table 1, but there's no comprehensive comparison between participants and non-participants. Cases' disease severity is distinguished, potentially limiting misclassification. Exposure assessment lacks detail and validation. Not all legumes, such as soy, are included in the exposure assessment. Information about potential knowledge of the outcome before diet assessment is missing. The outcome is measured by a technician, likely independent from exposure assessments. Adjustments are made for sex, age (matching), and referral indication, but many other confounders are overlooked. The study investigates protopathic bias, wherein gallstone patients might reduce legume intake due to pre-existing gastrointestinal symptoms, potentially limiting spurious correlations. Effect modification and interaction for sex and case type are also explored.<br><br>The risk of bias linked to assessment of dietary exposure is unknown as the study solely assessed legume intake frequency without quantities or certain sources like soya products. |
| Tutunchi et al. (2021) [15] | √ | √ | √ | ÷ | ÷ | √ | √ | √ | ÷ | √ | √ | + | ? | √ | Authors conclusions: Legume consumption is not associated with NAFLD.<br><br>There appear to be some differences between cases and controls regarding factors that could act as significant confounders in the relationship. Adjustments are made for only some differences. Cases and controls originate from the same clinic, making them relatively comparable due to shared referral indications. Case ascertainment (outcome assessment) is conducted by a radiologist before exposure status is known. While some covariables are measured using validated methods, there's no information about the validation of their dietary assessment. The use of a 3-day diet record may not be optimal, particularly with outcomes that have a latency period. Adjusting for hormone therapy might have been beneficial. The study acknowledges potential sparse-data bias and notes limitations in inferring causality due to the case-control design. The wide confidence intervals suggest that a larger sample size could provide a more robust estimate. |

*Yes (√), No (÷), Unknown (?), High quality (++), Acceptable (+), Unacceptable (0). NAFLD, non-alcoholic fatty liver disease. FFQ, food frequency questionnaire. OR, odds ratio. CI, confidence interval.*

#### Cross-sectional studies

*Quality assessment of included cross-sectional studies with Appraisal of Cross-sectional Studies (AXIS) tool.* [16]

| | 1. Were the aims/objectives of the study clear? | 2. Was the study design appropriate for the stated aim(s)? | 3. Was the sample size justified? | 4. Was the target/reference population clearly defined? | 5. Was the sample frame taken from an appropriate population base that closely represented the target/reference population under investigation? | 6. Was the selection process likely to select subjects/participants that were representative of the target/reference population under investigation? | 7. Were measures undertaken to address and categorize non-responders? | 8. Were the risk factor and outcome variables measured appropriate to the aims of the study? | 9. Were the risk factor and outcome variables measured correctly using instruments/measurements that had been trialled, piloted, or published previously? | 10. Is it clear what was used to determine statistical significance and/or precision estimates? (e.g, p-values, confidence intervals) | 11. Were the methods (including statistical methods) sufficiently described to enable repetition? | 12. Were the basic data adequately described? | 13. Does the response rate raise concerns about non-response bias? | 14. If appropriate, was information about non-responders described? | 15. Were the results internally consistent? | 16. Were the results presented for all the analyses described in the methods? | 17. Were the authors' discussions and conclusions justified by the results? | 18. Were the limitations of the study discussed? | 19. Were there any funding sources or conflicts of interest that may affect the authors' interpretation of the results? | 20. Was ethical approval or consent of participants attained? | Notes. |
|---|---|---|---|---|---|---|---|---|---|---|---|---|---|---|---|---|---|---|---|---|---|
| Baratta et al. (2017) [17] | √ | √ | ÷ | √ | √ | √ | ÷ | √ | √ | √ | √ | √ | ÷ | ÷ | √ | √ | √ | √ | √ | √ | Author conclusions: Consuming legumes were only associated with lower risk of NAFLD before adjustment. After adjusting for confounders, the association was nonsignificant.<br><br>The absence of details regarding participation rates and sample size calculations raises uncertainties about the recruitment process. It's unclear if all eligible individuals were invited to participate or if the selection was limited. They could have benefitted from a flow chart of participants.<br><br>While the study aims to investigate the link between NAFLD and MedDiet in patients with cardiometabolic risk factors, there's a lack of information about the source population from which the sample was drawn, creating ambiguity regarding the sample's representativeness. The study employed reliable and validated methods for measuring exposure, outcome, and covariables. It reported confidence intervals and p-values set at p<5%, and covariable adjustments were well-described in the statistical section. However, the study's presentation of participant data in Table 1 shows inconsistencies in adherence group sizes, suggesting a potential benefit from a more balanced range in the diet score to create more equally sized groups. Selection is not well described, and this may be associated with selection bias which could bias the estimate of an association towards the null or minimize the power of the study. Overall quality: Low |
| Bullón-Vela et al. (2019) [18] | √ | √ | √ | √ | √ | √ | ÷ | √ | √ | √ | √ | √ | √ | ÷ | √ | √ | √ | √ | √ | √ | Author conclusions: Legume consumption was inversely associated with NAFLD in individuals with MetS.<br>The methods are well considered, and the article presents the most important points necessary to assess the quality of the evidence. The article generally lacks information on non-participants, who were not described nor compared to participants. Overall quality: Moderate |
| Chan et al. (2015) [19] | √ | √ | ÷ | √ | √ | √ | ? | √ | ÷ | √ | √ | √ | ÷ | ÷ | √ | √ | √ | √ | √ | √ | Author conclusions: Higher consumption of legumes was associated with a reduced prevalence of NAFLD in Hong Kong Chinese<br>They don't justify the inclusion of 3000 individuals of which only 1/3 participate. With substantial dropout, information about non-responders is crucial for generalizing the study's findings to the broader population. They claim no major differences in baseline characteristics between included and excluded subjects without providing data. Internal validation is weak; the FFQ covers only the last 7 days, and spot urine measurements lack consistency and 24-hour estimates. Sodium and potassium calculations were lower than anticipated, affecting reliability, especially for nutrients not in standard portions. The use of reference portions for estimating usual intake is noted but lacks accuracy for certain nutrients like sodium. Participants knowing both exposure and outcome at baseline could lead to biased dropout, likely underestimating the link between diet and NAFLD. Internal validity is compromised due to questionable validation methods, raising concerns for bias. Overall quality: Very low |
| Chiu et al. (2018) [20] | √ | √ | √ | √ | √ | ? | ? | √ | √ | √ | √ | √ | ? | ÷ | √ | ? | ? | √ | √ | √ | Author conclusions: Choosing soy over meat or fish may prevent fatty liver.<br>The study utilized cohort data, including all available participants without specific reference to the Tzu Chi Health Study. Excluded individuals are presented, but there's a lack of detailed information or comparison with the full participant group. Physical activity, an important factor in NAFLD, was adjusted for but not displayed in the results. The authors claim it didn't impact the outcomes, yet these results aren't shown, making it challenging to assess potential confounding due to physical activity. The study methodology is generally good if the study population is sampled from a closed and well-defined cohort. However, the lack of references to more information on the Tzu Chi Health Study raises questions about the source population, posing risks of selection bias. In this very health-oriented population, it's likely there's an underrepresentation of individuals |

| | | | | | | | | | | | | | | | | | | | | | |
|---|---|---|---|---|---|---|---|---|---|---|---|---|---|---|---|---|---|---|---|---|---|
| | | | | | | | | | | | | | | | | | | | | | with unhealthy habits like smoking or alcohol intake. Consequently, any true associations might be stronger compared to the observed in this study. Overall quality: Moderate |
| Mirizzi et al. (2019) [21] | √ | √ | √ | √ | √ | √ | ? | √ | √ | √ | √ | √ | √ | ÷ | ? | √ | √ | √ | √ | √ | Author conclusions: The Mediterranean diet's emphasis on fresh foods from non-intensive farming and high legume intake appears beneficial for individuals with NAFLD.<br>This study provides a baseline assessment of an RCT, indicating a well-founded approach due to the controlled nature of RCTs in general. Some details regarding non-responders and potential imputations could enhance understanding of selection issues, yet these might not significantly alter the observed associations. There's a mention of excluded participants and missing data in baseline tables without a thorough comparison between full participants and non-participants. Despite discrepancies in participant numbers (Table 1 "Fatty liver index"), the study seems well-established. Overall quality: Moderate |
| Tseng et al. (2000) [22] | √ | √ | √ | √ | √ | √ | ÷ | √ | √ | √ | √ | √ | √ | ÷ | √ | ÷ | ÷ | √ | √ | √ | Author conclusions: Bean consumption was inversely associated with GBD among men (nonsignificant). and positively associated with GBD among cases unaware of disease status among women.<br>The study's external validity appears reasonable, benefiting from a large sample of a specific population. Utilizing NHANES as a foundation adds strength to the study due to its extensive and diverse representation. Exposure and outcomes are measured effectively, with tools adapted for the targeted population. However, there's a lack of comparison between responders and non-responders. The study describes sample selection well, particularly oversampling Mexican Americans, a group significant for the research. The funding from the National Institutes of Health adds credibility. The study presents imputation for missing responses and acknowledges limitations in dietary data collection and potential changes in consumption patterns over time. The absence of detailed presentation for multivariable analyses and ambiguous discussion regarding statistically insignificant associations could be improved for clarity. Overall quality: Low |
| Unisa et al. (2010) [23] | √ | √ | √ | √ | √ | √ | ÷ | ? | ÷ | √ | ÷ | √ | √ | ÷ | ÷ | √ | √ | ÷ | √ | ÷ | Authors conclusions: Chickpea consumption habits was associated with greater risk of GBD.<br>They've diligently undertaken recruitment efforts to ensure a broadly representative sample, aiming to minimize any biases in participant selection. However, while they state that subsequent analyses weren't affected by non-response, they don't provide any insights into the characteristics of those who didn't respond. The description of diet assessment lacks depth and fails to include details on validation or reliability, which leaves a gap in understanding. Similarly, the description of adjustment levels and exposure variables falls short. The methods section doesn't sufficiently clarify the adjustment levels, leaving uncertainty about whether the components in table 3 were mutually adjusted in the OR estimates. There's a notable absence of information about non-responders, which could impact the interpretation of the findings. Moreover, their logistic regression in table 3 excludes some participants, and discrepancies arise in the count of men and women compared to the total participant number, raising concerns about data accuracy. Crucially, they omitted any discussion of limitations and failed to document ethical approval, two critical aspects in research appraisal and transparency. The method of the study is poorly articulated; the only adequately described aspect is the robust sampling strategy. However, the exposures and risk factors lack sufficient description. Additionally, the levels of adjustment in the analyses are not detailed in the methodology section. Overall quality: Very low |
| Vijay et al. (2022) [24] | √ | √ | √ | √ | √ | √ | ÷ | √ | √ | √ | √ | √ | √ | ÷ | √ | √ | √ | √ | √ | √ | Authors conclusions: Dried legumes and pulses reduced OR for NAFLD with approx. 50%.<br><br>The participants were selected from the Trivandrum NAFLD cohort, representing a randomly sampled group from the entire Trivandrum population, ensuring a representative background. However, the study lacks details about non-responders, though their limited number may not significantly impact selection bias or generalizability. The exposure, outcomes, and covariates are well-validated and thoroughly described in the methodology. The study demonstrates clear methodology and comprehensive reporting of results, enhancing its credibility and accessibility to readers. A well-described study with thoughtful methodological considerations, despite the absence of information on non-responders. Overall quality: Moderate |
| Watzinger at al. (2020) [25] | √ | √ | √ | √ | ? | ? | ÷ | √ | √ | √ | √ | √ | √ | ÷ | √ | √ | √ | √ | √ | √ | Author conclusions: Legume intake was inversely correlated with liver fat content, but no significant association was found between legume consumption and OR for NAFLD.<br>Participants recruited through university hospital flyers might limit generalizability as they were there for specific health-related reasons, impacting participant selection. Non-responders remain undescribed. However, the article provides well-described tables, comprehensive indexing of analyses, and employs validated methods for assessing the exposure, outcome, and covariables. It highlights the inability to draw temporal relationships due to the cross-sectional design and acknowledges the small, homogeneous sample of metabolically healthy overweight individuals. Participant selection might be biased due to recruitment location, and non-responders are not detailed. Overall quality: Low |
| Yabe et al. (2021) [26] | √ | ? | ÷ | √ | √ | √ | ? | ? | ÷ | √ | ? | √ | √ | ÷ | ? | ? | √ | √ | √ | √ | Author conclusions: Higher soybean consumption was associated with lower odds for NAFLD.<br><br>The study, being cross-sectional, lacks the ability to explore temporal progression. Sample size information is absent. Participants are sourced from health check-ups, likely representative given the insurance-based Japanese healthcare system. Exclusion criteria for those omitted are described. Exposure and covariate assessment were not validated. Outcome evaluation methods are validated. However, the study doesn't adjust for metabolic criteria leading to potential confounding. P-values are provided, and Bonferroni correction is applied for multiple comparisons. The methods were somewhat detailed, yet adjustment levels weren't outlined until the results section. Reference groups in logistic regression (Figure 5) weren't clearly explained. No comparison was made between participants and non-participants, and there's uncertainty about the presentation of all results, given the brief methods and discrepancies in adjustment levels between sections.<br><br>Overall, the study struggles with insufficient method description and presents results without corresponding methodological details. Overall quality: Moderate |
| Zhang et al. (2020) [27] | √ | √ | √ | ? | ? | √ | ? | √ | √ | √ | √ | √ | √ | ÷ | √ | √ | √ | √ | √ | √ | Author conclusion: Authors observed an inverse association between habitual soy food intake and NAFLD.<br><br>The study uses cross-sectional data from a Chinese cohort. The participant selection originates from a broader Chinese cohort, referenced unclearly, lacking a comprehensive description or a protocol reference. The cohort aims to represent the population of Tianjin Province. Limited flowchart depiction for excluded participants is present. Methods for exposure, outcomes, and risk factors are well-documented, previously utilized, and validated. They employ p-values and conduct numerous sensitivity analyses to assess primary analysis robustness, all detailed in the methods. However, there's no comparison between participants and non-participants. The analyses and results are well-described. Overall, the |

| | | | | | | | | | | | | | | | | | | | | | |
|---|---|---|---|---|---|---|---|---|---|---|---|---|---|---|---|---|---|---|---|---|---|
| | | | | | | | | | | | | | | | | | | | | | study lacks clarity in defining the source population, referring to various articles defining their own populations, requiring a more comprehensive study description or a protocol reference. Nonetheless, the study offers a well-described and replicable methodology. Overall quality: High |

*Yes (√), No (÷), Unknown (?). The answer Yes (√), marked with green, indicates no issues with response bias and funding or conflicts of interests for assessment point 13 and 19 respectively. The answer No (÷), marked with red, indicates issues with response bias and funding or conflicts of interests for assessment point 13 and 19 respectively. NAFLD, non-alcoholic fatty liver disease. MedDiet, Mediterranean diet. MetS, metabolic syndrome. FFQ, food frequency questionnaire. RCT, randomized controlled trial. GBD, gallbladder disease*

#### *Systematic reviews*

*Critical appraisal of included systematic reviews using A MeaSurement Tool to Assess systematic Reviews (AMSTAR-2).* [28]

| | 1. Question and inclusion | 2. Protocol | 3. Study design | 4. Comprehensive search | 5. Study selection | 6. Data extraction | 7. Excluded studies justification | 8. Included studies details | 9. Risk of bias | 10. Funding sources | 11. Statistical methods | 12. Risk of bias on meta-analysis | 13. Risk of bias in individual studies | 14. Explanation for heterogeneity | 15. Publication bias | 16. Conflict of interest | Notes. |
|---|---|---|---|---|---|---|---|---|---|---|---|---|---|---|---|---|---|
| He et al. (2020) [29] | √ | √ | ÷ | ? | √ | √ | √ | ? | √ | ÷ | √ | √ | √ | √ | ÷ | √ | Author conclusions: Most foods examined in the meta-analysis might not have a substantial impact on the likelihood of NAFLD.<br><br>The rationale for choosing observational studies isn't adequately explained. The search terms utilized were specified, and the selection process involved a duplicate screening procedure, resolving discrepancies with a third reviewer when necessary. Although they provided a flowchart summarizing the selection process, detailed explanations for exclusions in each potentially relevant study were lacking. Supplementary Table 1 and 2 offered an overview of included studies, but comprehensive descriptions were missing. The ROBINS-I tool was employed for bias assessment (Supplementary Table 3). Additionally, they conducted sensitivity analysis for low-bias studies and attempted publication bias assessment using Egger's tests and funnel plots when enough studies were available. Of included studies relevant for this scoping review, three had low bias and one had moderate. The meta-analysis exhibited homogeneity among three cross-sectional studies. However, there wasn't enough literature for a statistical assessment of publication bias. While funding details were absent, conflicts of interest were disclosed. Overall confidence in results: Moderate |

*Yes (√), No (÷), Partly yes (?). NAFLD, non-alcoholic fatty liver disease. ROBINS-I, risk of bias in non-randomised studies of interventions.*

### Appendix VI: Characteristics of included studies

*Articles focusing on non-alcoholic fatty liver disease (NAFLD)*

| Author, year, country | Sample characteristics | Legume exposure | Outcomes | Key findings |
|---|---|---|---|---|
| Cohort studies | | | | |
| Zhang et al. (2023) [5], China | Tianjin Chronic Low-grade Systemic Inflammation and Health Cohort (TCLSIH) n=14,541, 59.9% females<br><br>Median age 35.9 (IQR[a]: 30.3-45.5) years.<br><br>Guangzhou Nutrition and Health Study (GNHS) n=1297, 69.5% females. Median age 59.7 (IQR: 56.5-64.1) years. | Both cohorts standardized food component intakes to 25 g legumes per serving. Substitution food serving sizes were eggs 50 g; unprocessed red meat 50 g; processed meat 30 g; poultry 50 g; fish 50 g; nuts 10 g; legumes 25 g; whole grains 50 g | Risk of NAFLD[b] | HR[c] (95% CI[d]) of NAFLD when substituting 1 serving of legumes for eggs: 1.01 (0.93; 1.09), processed meats: 0.86 (0.74;1.00), unprocessed meats: 1.02 (0.96, 1.09), poultry: 0.92 (0.86, 1.00), fish: 0.99 (0.93, 1.05) in TCLSIH.<br><br>HR (95%CI) of NAFLD when substituting 1 serving of legumes for eggs: 0.66 (0.36, 1.24), meats: 1.16 (0.99, 1.37), poultry: 0.35 (0.18, 0.69), fish: 1.09 (0.92, 1.30) in the GNHS. |
| Case control studies | | | | |
| Bahrami et al. (2019) [6], Iran | Iranian adults from Teheran. 196 cases of NAFLD., 803 controls matched for age and sex.<br><br>Mean age 41.9±11.1 years, 51% females. | Weekly servings of legumes | Odds of NAFLD | OR[e] (95% CI) for NAFLD per increase of 1 serving of legumes weekly was 0.09 (0.030; 0.024) for total legumes, 0.58 (0.45; 0.75) for lentils, and 0.32 (0.16; 0.64) for beans after adjusting for energy intake (kcal/day), BMI, physical activity, prevalence of dyslipidaemia and diabetes, smoking status, and dietary intake (g/day) of fats, protein, carbohydrate, fruits, vegetables, and whole grains.<br><br>OR (95% CI) for NAFLD per increase of 1 serving of legumes weekly was 0.74 (0.64; 0.84) for total legumes, |

| | | | | |
|---|---|---|---|---|
| | | | | 0.61 (0.46; 0.78) for lentils, and 0.35 (0.17; 0.74) for beans after adjusting for energy intake (kcal/day), BMI, physical activity, prevalence of dyslipidaemia and diabetes, smoking status, and dietary intake (g/day) of red and processed meat and high-fat dairy. |
| Giraldi et al. (2020) [7], Italy | Italian adults from Rome. 371 cases of NAFLD, 444 controls, matching criteria unknown. Female cases: 32.8%, controls: 41.2%.<br><br>Mean age (SD) cases: 59 (16.0), controls: 45 (14.4). | Legume consumption was categorized in two groups: low or high. | Odds of NAFLD | Highest vs. lowest legume consumption was associated with lower OR for NAFLD (OR: 0.62, 95% CI: 0.38-0.99). |
| Han et al. (2014) [8]. | Korean adults. 169 cases of NAFLD, 179 controls, matching criteria unknown. 166 men, 182 women.<br><br>Age 20-69 years. | Daily legume consumption scored in tertiles | Odds for NAFLD | OR (95% CI) for NAFLD when comparing highest with lowest tertile of bean intake was for men 1.50 (0.59; 3.84) and women 1.13 (0.44; 2.89) after adjusting for age, employment, educational level, physical activity, smoking status, and energy intake. |
| Hao et al. (2021) [9], China | Chinese adults. 2602 cases of NAFLD, 1447 controls, matched on genotype.<br><br>Mean (SD) age for cases: 52.98 (8.15), controls: 53.06 (7.89). Female participants 69-71%. | Weekly consumption frequency of beans | Odds of NAFLD | Compared to seldom consumption those consuming beans 1-4 times weekly had an OR for NAFLD of 0.710 (95% CI: 0.607; 0.831), while those consuming beans 5-7 times weekly had an OR of 0.701 (95% CI: 0.576; 0.853).<br><br>When stratifying on methylene tetrahydrofolate reductase, those with CC genotype consuming beans 1-4 times weekly compared to seldom had an OR for NAFLD of 0.612 (95% CI: 0.443; 0.847), and those consuming beans 5-7 times weekly compared to seldom had an OR of 0.807 (95% CI: 0.541; 1.204). Among those with the CT or TT genotype consuming beans 1-4 times weekly compared to seldom had an OR for NAFLD of 0.745 (95% CI: 0.622; 0.892), and |

| | | | | |
|---|---|---|---|---|
| | | | | those consuming beans 5-7 times weekly compared to seldom had an OR of 0.670 (95% CI: 0.535; 0.839). |
| Tutunchi et al. (2021) [15], Iran | Iranian adults. 105 cases of NAFLD, 105 controls, matched on sex and age.<br><br>57.2% females, mean age cases: 45.6 years, controls: 45.4 years. | Legume intake g/day tertiles | Odds of NAFLD | When comparing tertiles of legume intake, the OR (95% CI) for NAFLD was 0.68 (0.21; 1.32) for second compared the first tertile, and 0.74 (0.61; 1.67) for third compared to first tertile when adjusting for age, sex, education, physical activity, BMI, and waist circumference. |
| Cross-sectional studies | | | | |
| Baratta et al. (2017) [17], Italy | Italian adults. 584 patients with one or more cardiovascular risk factors: T2DM[f], hypertension, overweight/obesity, dyslipidaemia, MetS[g]. Mean (SD) age 56.2 (12.4) years, 38.2% females. | Adherence to the Mediterranean diet was scored from 0 to 9. More than two servings of legumes weekly gave 1 point. | Odds of NAFLD | OR (95% CI) for NAFLD when consuming ≥ 2 servings legumes compared to fewer was 0.548 (0.355; 0.847) in an unadjusted model and 0.675 (0.370; 1.230) after adjusting for sex, age, waist circumference (>102 cm for men, >88 cm for women), triglycerides ≥150 mg/dl, hypertension, use of statins, alanine aminotransferase levels, previous major adverse cardiovascular and cerebrovascular events, type 2 diabetes, and adherence to the Mediterranean diet. |
| Bullón-Vela et al. (2019) [18], Spain | Spanish adults. 328 individuals with Mets from the PREDIMED-Plus[h] trial. 45.1% females, mean age 65.8 years. | Legume consumption g/day in tertiles | HSI in tertiles as an indication for fat content in liver | Linear regression analysis demonstrated a negative relationship between HSI[i] and legume consumption ($R^2$ adjusted = 0.027, P=0.002, beta = -0.093). An inverse linear association between legume intake (g/d) and the highest tertile of HSI was observed (β= -0.093, p=0.002).<br><br>Multivariate analysis of relative risk ratio (RRR) between legume consumption tertiles and HSI tertiles showed that highest compared to lowest legume intake among those in second HSI tertile was associated with a RRR of 0.81 (95% CI: 0.42; 1.59) when adjusting for age, energy intake, alcohol consumption, and smoking status. Further adjusting for triacylglycerols and physical activity resulted in RRR of 0.74 (95% CI: 0.37; 1.46). Highest compared to lowest |

| | | | | |
|---|---|---|---|---|
| | | | | legume intake among those in third HSI tertile was associated with a RRR of 0.54 (95% CI: 0.27; 1.06) when adjusting for age, energy intake, alcohol consumption, and smoking status. Further adjusting for triacylglycerols and physical activity resulted in RRR of 0.48 (95% CI: 0.24; 0.97). |
| Chan et al (2015) [19], China | Chinese adults. 797 participants from a NAFLD screening program in Hong Kong, China. Mean (SD) age 48.1 (10.6) years. 41.7% males. | Daily soy and soy product intakes were grouped in tertiles. | Odds of NAFLD | OR (95% CI) for NAFLD among those with highest compared to lowest intake of soy and soy products was 0.93 (0.63; 1.38) in age- and sex-adjusted analysis. When further adjusting for BMI, smoking status, alcohol consumption status, central obesity, triglyceride >1.7 mmol/l, reduced HDL-cholesterol, hypertension, impaired fasting glucose or diabetes, genotype, and energy intake, highest compared to lowest intake of soy and soy products resulted in an OR for NAFLD of 0.86 (95% CI: 0.52; 1.42). |
| Chiu et al. (2018) [20], Taiwan | Taiwanese adults. 4625 participants from the Tzu Chi Health Study.<br><br>Vegetarians: mean age 54 years, 59% females. Non-vegetarians: mean age 55 years, 78% females. | Servings of protein from legumes substituted for a serving of protein from other sources | Odds of NAFLD | Substituting one serving of meat for one serving of soy resulted in an OR of NAFLD of 0.96 (95% CI: 0.91; 1.03) when adjusting for age, sex, education, smoking history, alcohol consumption history, energy intake, and vegetarian diet. When further adjusting for BMI, the OR for NAFLD was 0.95 (95% CI: 0.88; 1.02).<br><br>Figure 2 indicates that substituting meat or fish for soy increases the OR for NAFLD with 12-13% (lower soy causing increased OR, estimates not presented in text). |
| Mirizzi et al. (2019) [21], Italy | Italian adults. 136 participants with NAFLD from the NUTRIATT-RCT[30,j]. Mean age (SD) was 49.58 (10.18) years. 58% males. | Legume consumption g/day | Odds of NAFLD | When comparing individuals with severe NAFLD to those with moderate, the OR (95% CI) for consumption of soy milk was 0.98 (0.97; 0.99), for chickpeas was 0.71 (0.56; 0.92), and for dried peas was 0.69 (0.51; 0.94) when adjusting for age, sex, and energy intake. When further adjusting for all other food groups (chocolate, winter ice- |

| | | | | |
|---|---|---|---|---|
| | | | | cream, apricots, pears, soy milk, legume-rice, chickpeas, dried peas, local aged cheeses, industrial aged cheeses, white bread, sweet milk, regular ice-cream, French fries, fats, and rabbit meat) apart from the one assessed, the OR (95% CI) for consumption of soy milk was 0.99 (0.97; 1.02), for chickpeas was 0.57 (0.34; 0.97), and for dried peas was 0.78 (0.44; 1.39). |
| Vijay et al. (2022) [24], UK | South Indian adults. 1966 individuals comprising 993 NAFLD cases and 973 controls.<br><br>Cases: mean age 48.16 years, 54.4% females. Controls: mean age 45.90 years, 67.5% females. | Consumption of legumes per kg of bodyweight per day | Degree of liver fibrosis measured as liver stiffness > 8.4 kPa | The linear association between weight-adjusted mean intakes (mean g/kg/day ± SD) of legumes and liver fibrosis indicated inverse associations for pulses and legumes (β: -0.015, SE: 0.006), dried pulses and legumes (β: -0.016, SE: 0.008), and fresh pulses and legumes (β: -0.010, SE: 0.004). None of the associations were statistically significant.<br><br>OR for NAFLD was inversely associated with intake of dried pulses and legumes (OR: 0.43, 95% CI: 0.21; 0.61). |
| Watzinger et al. (2020) [25], Germany | Germen adults. 136 participants from the HELENA-trial[31,k].<br><br>Without NAFLD: 57.8 %, females, mean age 49.8 years. With NAFLD: 44.4% females, mean age 50.1 years. | Legume intake g/day | Odds of NAFLD | OR for NAFLD comparing the lowest and highest quartiles of legume consumption was 1.70 (95% CI: 0.56; 5.17) when adjusting for sex, age, waist circumference, and energy intake. When further adjusting for the ratio of energy intake/total energy expenditure, the OR for NAFLD was 1.78 (95% CI: 0.57; 5.55). |
| Yabe et al. (2021) [26], Japan | Japanese adults. 349 patients who visited an outpatient clinic for lifestyle related diseases.<br><br>248 had NAFLD (118 men, 130 women, mean (SD) age 55.8 (15.1) years). 101 did not have NAFLD | Weekly frequency of soy and soybean consumption | Liver fibrosis measured with Fibro-AST[l] score | Mean frequency consumption of soybeans and soybean products varied significantly between the three Fibro-AST groups (p=0.005), with higher intakes among those with lower liver fibrosis scores.<br><br>Individuals who consumed soybeans or soybean products ≥ 4 times/week had an OR for liver fibrosis of 0.57 (95% CI: 0.27; 1.19). Comparison group not defined. |

| | | | | |
|---|---|---|---|---|
| | (29 men, 72 women, mean (SD) age 58.4 (18.2) years). | | | |
| Zhang et al. (2020) [27], China | Northern Chinese adults. 24,622 participants from TCLSIH.<br><br>Individuals without NAFLD: 58.8% females, mean age 38.4 years, Individuals with NAFLD: 27.7% females, mean age 42.4 years. | Weekly soy food consumption frequency | Odds of NAFLD | OR for NAFLD for those consuming soy foods ≥4 times weekly compared to less than once was 0.74 (95% CI: 0.64; 0.86) after adjusting for age, sex, BMI, energy intake, smoking status, alcohol drinking status, educational level, occupation, household income, physical activity, hypertension, hyperlipidaemia, diabetes, family history of cardiovascular disease, hypertension, hyperlipidaemia, and diabetes, intake of eicosapentaenoic acid (EPA) and docosahexaenoic acid (DHA), total protein intake, and total carbohydrate intake. When further adjusting for the Chinese Healthy Eating index (CHEI) excluding the soy food component in a fully adjusted model, the OR for NAFLD was 0.75 (95% CI: 0.65; 0.87).<br><br>Stratified analyses showed significant inverse associations between soy food intake ≥4 times/wk compared to <1 time/wk and NAFLD in men (OR: 0.73, 95% CI: 0.61; 0.87), age < 50 years (OR: 0.70, 95% CI: 0.59; 0.84), or BMI ≥ 24 kg/m2 (OR: 0.76, 95% CI: 0.65; 0.89) in the fully adjusted model.<br><br>OR for NAFLD based on energy-adjusted daily increase in soy food intake by 10 g/1000 kcal was 0.94 (95% CI: 0.91; 0.97) in the fully adjusted model.<br><br>When excluding individuals with hypertension, hyperlipidaemia, and diabetes (n=11,677) the OR for NAFLD for those consuming soy foods ≥4 times weekly compared to less than once was 0.72 (95% CI: 0.55, 0.95) in the fully adjusted model. |

| | | | | |
|---|---|---|---|---|
| Systematic reviews | | | | |
| He et al. 2020 [29], China | Of 7892 articles 24 observational studies were included. Of these, 4 investigated legume consumption and NAFLD risk.<br><br>Inclusion: Adult participants, observational studies investigating food groups and likelihood of validated NAFLD diagnosis.<br><br>Exclusion: Animal studies, adolescents, pregnant women, present hepatitis B or C, HIV, or cancer, consumption of alcohol >20 g/day for females and >30 g/day for males, and other factors causing hepatic steatosis. | The review included food groups as exposures with legume composing one food group. | The outcome was likelihood of valid diagnosis of NAFLD. | The pooled results of three homogenous cross-sectional studies in the meta-analysis revealed no substantial association between legume consumption and NAFLD (OR: 0.943, 95% CI: 0.877; 1.014). The case-control study found a negative association between legume intake and NAFLD (OR: 0.730, 95% CI: 0.637; 0.836).<br><br>The bias assessment indicated low risk of bias in three studies and moderate risk in one. |
| Narrative reviews | | | | |
| Mega et al. (2021) [32], Italy | Mixed populations, healthy adults and NAFLD patients | Legume consumption | NAFLD likelihood | In animals, legumes upregulate genes linked to beta-oxidation and acetyl-CoA degradation while downregulating those involved in glycolysis and lipogenesis. A RCT of 42 premenopausal women with central obesity showed that a hypocaloric diet enriched in legumes led to significant decreases in AST[m] and ALT[n] blood levels after 6 weeks compared to a hypocaloric diet without legumes. A case-control study highlighted a significant association between lower NAFLD risk and higher legume intake (OR of NAFLD for total legumes 0.73; 95% CI 0.64; 0.84, lentils 0.73; 95% CI 0.64; 0.84%, and beans 0.35; 95% CI 0.17; |

| | | | | |
|---|---|---|---|---|
| | | | | 0.74). These estimates are incorrectly reported compared to the primary study by Bahrami et al 2019 [6].<br><br>A meta-analysis by He et al. [29] of three cross-sectional studies found no significant link between legume consumption and NAFLD likelihood.<br><br>Soybeans appear to have a distinct role in NAFLD prevention. Substituting soy for meat or fish showed a 12–13% increased risk of fatty liver disease in a large Chinese cross-sectional study by Chiu et al [20].<br><br>The protective effect is likely attributed, at least in part, to the high β-conglycinin (7S globulin) content, known to downregulate hepatic expression of the PPARγ-2 gene in animal models. |

[a]IQR, interquartile range. [b]NAFLD, non-alcoholic fatty liver disease. [c]HR, hazard ratio. [d]CI, confidence interval. [e]OR, odds ratio. [f]T2DM, type 2 diabetes mellitus. [g]MetS, metabolic syndrome. [h]PREDIMED-Plus, Prevención con Dieta Mediterránea-Plus. [i]HSI, hepatic steatosis index. [j]NUTRIATT-RCT, nutrition and activity randomized controlled trial. [k]HELENA-trial, Healthy nutrition and energy restriction as cancer prevention strategies: a randomized controlled trial. [l]Fibro-AST, fibrosis-aspartate amino transferase. [m]AST, aspartate amino transferase. [n]ALT, alanine amino transferase.

*Articles focusing on gallbladder diseases (GBD)*

| Author, year, country | Sample characteristics | Legume exposure | Outcomes | Key findings |
|---|---|---|---|---|
| Cohort studies | | | | |
| Barré et al. (2017) [2], France | French adult women. 64,052 women from the Etude Epidémiologique auprès de Femmes de la Mutuelle Générale de l'Education Nationale cohort[3]. | Legume consumption g/day | Cholecystectomy as proxy for gallstones | Incidence rate of cholecystectomy 268.7/100,000 person-years (95% CI[a]: 258.7; 278.7).<br><br>Those with highest consumption of legumes (≥ 27.9 g/day) had a HR[b] of 0.73 (95% CI: 0.65; 0.82) for incident cholecystectomy compared to those with lowest legume |

| | | | | |
|---|---|---|---|---|
| | Mean age was 53.7 years for those having cholecystectomy and 52.6 years for those not having cholecystectomy. | | | consumption (0 g/day) when adjusting for age, educational level, BMI, use of oral contraceptives, menopausal hormone therapy, smoking status, energy intake excluding alcohol, alcohol, physical activity, number of livebirths, diabetes, and cholesterol-lowering drug. |
| Tsai et al. (2006) [4], USA | American adult women. 77,090 women from the Nurses' Health Study. Participants were aged 37-64 at recruitment in 1984. | Legume intake was divided into quintiles of weekly servings ranging <0.5 to >2. | Self-reported cholecystectomy | Those consuming > 2 compared to < 0.5 servings of legumes weekly had a HR for cholecystectomy of 1.02 (95% CI: 0.94; 1.11) when adjusting for age and pack-years of smoking. When further adjusting for time period of data collection, BMI, weight change the previous 2 years, parity, oral contraceptive use, hormone replacement therapy, physical activity, history of diabetes, energy intake, alcohol consumption, coffee consumption, and use of thiazide diuretics and nonsteroidal anti-inflammatory drugs, the HR for cholecystectomy was 1.02 (95% CI: 0.94; 1.12).<br><br>Sub analysis adjusting for macronutrients, fat types, and dietary fibre did not alter the results of main analyses. |
| Case-control studies | | | | |
| Jayanthi et al. (2005) [10], India | Indian adults. 346 gallstone cases and 346 sex and age matched controls, 51.4% females, mean (SD) age cases: 51 (14.1) years, controls: 50.3 (14.8) years. | Consumption frequency of Tamarind bean paste | Odds of gallstone formation | OR[c] for gallstone formation when using tamarind (bean paste) 3 or more times weekly was in univariate analysis 1.76 (95% CI: 1.05; 2.96). In multi-variate adjusted analysis the OR was 1.79 (95% CI: 1.09; 2.93) when consuming tamarind ≥3 times weekly compared to <3 times weekly. Adjustment variables are not mentioned. |
| Parveen et al (2019) [11], Pakistan | Pakistani adults. 100 gallstone cases and 100 sex- and age-matched controls. Male to female ratio was 1:2. | Weekly legume consumption | Odds of gallstones | OR (95% CI) for gallstones when consuming legumes once weekly was 3.37 (95% CI: 1.20; 9.60) for peas, 3.69 (95% CI: 0.67; 22.84) for mung beans (vigna radiate), and 0.57 (95% CI: 0.15; 2.08) for masoor (lens culinaris) compared to consuming mixed pulses. |

| | | | | |
|---|---|---|---|---|
| Pastides et al. (1990) [12], Greece | Greek adult women. 84 women with gallstones and 171 controls.<br><br>Mean (SD) age of cases was 54.8 (11.48) years and controls 54.3 (10.77) years. | Monthly legume consumption | Linear trend for gallstone | There was no significant association found between pulses/beans and the risk of gallstone in the linear trend analysis (chi-squared: 2.97, p=0.085). As a result, pulses/beans were excluded from the multivariate analysis. |
| Rasheed et al. (2020) [13], Saudi Arabia | Saudi Arabian adult women. 157 cases of gallstones and 175 healthy controls from the Qassim region. Participants were 20-50 years old. | Legume consumption | Gallstones | Legume consumption was significantly higher among cases (43.3 %) than controls (33.3 %), p<0.05. |
| Thijs et al. (1990) [14], The Netherlands | Dutch adults. 204 gallstone cases and 615 age and sex matched controls. 67.7% cases were females, for controls it was 69.4%. Age ranged 30-76 for males and 20-76 for females. | Monthly legume intake frequency | Odds of gallstones | OR for gallstones was 0.40 (95% CI: 0.23; 0.87) among those who consumed legumes on ≥ 8 days monthly compared to < 1 day monthly. |
| Cross-sectional | | | | |
| Tseng et al. (2000) [22], USA | Mexican American adults. 4641 participants (2306 women, 2335 men) from the 1988-1994 NHANES III. Age ranged 20-74 years at inclusion. | Weekly consumption frequency of legumes | Prevalence odds of GBD[d] | Prevalence OR for GBD comparing legume consumption >1 time/day with < 1 time/wk was 1.05 (95% CI: 0.74; 1.51) for women and 0.59 (0.24; 1.44) for men after adjusting for age.<br><br>Age-adjusted prevalence OR for GBD among women unaware of GBD status comparing legume consumption >1 time/day with < 1 time/wk was 1.58 (1.02; 2.48). Results for men was not shown. |
| Unisa et al. (2010) [23], India | Indian adults. 6548 individuals (2625 males, 3923 females) underwent ultrasound examinations | Chickpea consumption habits | Odds of GBD | In total, GBD prevalence was 6.20%, and GST accounted for 4.15%. Among females, GBD prevalence was 1.7 times higher than in males. Symptomatic individuals had a greater GBD prevalence (7.12%) than asymptomatic individuals (2.99%). Symptomatic males had 2.5 times greater GBD |

| | | | | |
|---|---|---|---|---|
| | for GBD. All participants were ≥ 30 years old. | | | prevalence than asymptomatic males, and symptomatic females had 2.4 times greater prevalence than asymptomatic females.<br><br>Females exhibited 2.9 times greater gallstone prevalence than males. Gallstone prevalence was higher in symptomatic individuals (4.72%) than asymptomatic individuals (2.31%). Symptomatic individuals, both male and female, had 2.3 times greater GST prevalence than those without symptoms.<br><br>OR for GBD when consuming compared to not consuming chickpeas was 1.636 (95% CI: 1.261; 2.122). In stratified analyses, the OR for GBD when consuming compared to not consuming chickpeas was 2.546 (95% CI: 1.563;4.146) for men and 1.354 (95% CI: 0.981; 1.860) for women. |
| Narrative reviews | | | | |
| Di Ciaula et al. (2019) [33], Italy | Populations consuming large amounts of beans; Amerindians Mapuche and Pima Indians | Bean intake | Gallstone occurrence | Genetic factors significantly influence gallstone formation, especially when combined with dietary and metabolic triggers.<br><br>In Western countries, 75% of gallstones are cholesterol-based and associated with metabolic abnormalities linked to altered cholesterol homeostasis like obesity, T2DM[e], and MetS[f]. Overweight and obesity significantly increase the risk of gallstone formation and cholecystectomy.<br><br>Insulin levels and high fructose intake independently contribute to gallstone risk, while vitamin C regulates cholesterol homeostasis by converting biliary cholesterol into bile acids. |

| | | | | |
|---|---|---|---|---|
| Gaby (2009) [34], USA | Participants from Nurses' Health Study, Chileans, American Indians, and Dutch individuals. | The exposure across studies is legume consumption | GBD mainly focusing on gallstones. | Sugars, fats, and meats are mentioned as potential confounders often adjusted for in primary literature. Food allergies may inhibit gallbladder emptying by causing cholecystitis-like symptoms and could thus be potential confounders or modifiers of the association between foods and GBD. |
| Nutrition reviews (1989) [35], USA | Studies including healthy and patient populations who consume beans as a staple i.e., Pima Indians and Chileans. | Legume consumption | Altered cholesterol synthesis and gallstone risk | Consuming legumes has been linked to a higher occurrence of gallstones in both healthy individuals and patients. This increased prevalence, primarily seen in normal and obese subjects, is attributed to cholesterol supersaturation in the bile, which is linked to higher legume intake. |

[a]CI, confidence interval. [b]HR, hazard ratio. [c]OR, odds ratio. [d]GBD, gallbladder disease. [e]T2DM, type 2 diabetes mellitus. [f]MetS, metabolic syndrome.

#### Appendix VII: Preferred Reporting Items for Systematic reviews and Meta-Analyses extension for Scoping Reviews (PRISMA-ScR) Checklist[36]

| SECTION | ITEM | PRISMA-ScR CHECKLIST ITEM | REPORTED ON PAGE # |
|---|---|---|---|
| **TITLE** | | | |
| Title | 1 | Identify the report as a scoping review. | 1 |
| **ABSTRACT** | | | |
| Structured summary | 2 | Provide a structured summary that includes (as applicable): background, objectives, eligibility criteria, sources of evidence, charting methods, results, and conclusions that relate to the review questions and objectives. | 2-3 |
| **INTRODUCTION** | | | |
| Rationale | 3 | Describe the rationale for the review in the context of what is already known. Explain why the review questions/objectives lend themselves to a scoping review approach. | 4-6 |
| Objectives | 4 | Provide an explicit statement of the questions and objectives being addressed with reference to their key elements (e.g., population or participants, concepts, and context) or other relevant key elements used to conceptualize the review questions and/or objectives. | 6-8 |
| **METHODS** | | | |
| Protocol and registration | 5 | Indicate whether a review protocol exists; state if and where it can be accessed (e.g., a Web address); and if available, provide registration information, including the registration number. | 8 |
| Eligibility criteria | 6 | Specify characteristics of the sources of evidence used as eligibility criteria (e.g., years considered, language, and publication status), and provide a rationale. | 9 |
| Information sources* | 7 | Describe all information sources in the search (e.g., databases with dates of coverage and contact with authors to identify additional sources), as well as the date the most recent search was executed. | 8-9 |
| Search | 8 | Present the full electronic search strategy for at least 1 database, including any limits used, such that it could be repeated. | Appendix I-II |

| SECTION | ITEM | PRISMA-ScR CHECKLIST ITEM | REPORTED ON PAGE # |
|---|---|---|---|
| Selection of sources of evidence† | 9 | State the process for selecting sources of evidence (i.e., screening and eligibility) included in the scoping review. | 10 |
| Data charting process‡ | 10 | Describe the methods of charting data from the included sources of evidence (e.g., calibrated forms or forms that have been tested by the team before their use, and whether data charting was done independently or in duplicate) and any processes for obtaining and confirming data from investigators. | 10-12, Appendix IV |
| Data items | 11 | List and define all variables for which data were sought and any assumptions and simplifications made. | 7 |
| Critical appraisal of individual sources of evidence§ | 12 | If done, provide a rationale for conducting a critical appraisal of included sources of evidence; describe the methods used and how this information was used in any data synthesis (if appropriate). | 11-12 |
| Synthesis of results | 13 | Describe the methods of handling and summarizing the data that were charted. | 10-12 |
| **RESULTS** | | | |
| Selection of sources of evidence | 14 | Give numbers of sources of evidence screened, assessed for eligibility, and included in the review, with reasons for exclusions at each stage, ideally using a flow diagram. | 12, Figure 1, Appendix III |
| Characteristics of sources of evidence | 15 | For each source of evidence, present characteristics for which data were charted and provide the citations. | 13, Table 1 |
| Critical appraisal within sources of evidence | 16 | If done, present data on critical appraisal of included sources of evidence (see item 12). | 13-18, Appendix V |
| Results of individual sources of evidence | 17 | For each included source of evidence, present the relevant data that were charted that relate to the review questions and objectives. | 13-18, Appendix VI, Table 2, Table 3 |
| Synthesis of results | 18 | Summarize and/or present the charting results as they relate to the review questions and objectives. | 13-18, Appendix VI, Table 2, Table 3 |
| **DISCUSSION** | | | |

| SECTION | ITEM | PRISMA-ScR CHECKLIST ITEM | REPORTED ON PAGE # |
|---|---|---|---|
| Summary of evidence | 19 | Summarize the main results (including an overview of concepts, themes, and types of evidence available), link to the review questions and objectives, and consider the relevance to key groups. | 18-30 |
| Limitations | 20 | Discuss the limitations of the scoping review process. | 30 |
| Conclusions | 21 | Provide a general interpretation of the results with respect to the review questions and objectives, as well as potential implications and/or next steps. | 30-31 |
| **FUNDING** | | | |
| Funding | 22 | Describe sources of funding for the included sources of evidence, as well as sources of funding for the scoping review. Describe the role of the funders of the scoping review. | 31 |

JBI = Joanna Briggs Institute; PRISMA-ScR = Preferred Reporting Items for Systematic reviews and Meta-Analyses extension for Scoping Reviews.
* Where *sources of evidence* (see second footnote) are compiled from, such as bibliographic databases, social media platforms, and Web sites.
† A more inclusive/heterogeneous term used to account for the different types of evidence or data sources (e.g., quantitative and/or qualitative research, expert opinion, and policy documents) that may be eligible in a scoping review as opposed to only studies. This is not to be confused with *information sources* (see first footnote).
‡ The frameworks by Arksey and O'Malley[37] and Levac and colleagues[38] and the JBI guidance[39,40] refer to the process of data extraction in a scoping review as data charting.
§ The process of systematically examining research evidence to assess its validity, results, and relevance before using it to inform a decision. This term is used for items 12 and 19 instead of "risk of bias" (which is more applicable to systematic reviews of interventions) to include and acknowledge the various sources of evidence that may be used in a scoping review (e.g., quantitative and/or qualitative research, expert opinion, and policy document).
